# Identifying trial-relevant concepts of interest in HSP: insights from an international patient-voice study in over 600 individuals

**DOI:** 10.64898/2026.04.09.26350392

**Authors:** Mirlinda Ademi, Jonas Alex Morales Saute, Charlotte Dubec-Fleury, Julie Greenfield, Ruby Wallis, Chantal Gobeil, Lori Renna Linton, Andreas Nadke, Rita Horvath, Stephan Klebe, Filippo M. Santorelli, Atay Vural, Bart van de Warrenburg, Cynthia Gagnon, Matthis Synofzik, PROSPAX Consortium, Sophie Tezenas du Montcel, Rebecca Schüle

**Affiliations:** Division of Neurodegenerative Diseases and Movement Disorders, Department of Neurology, Heidelberg University Hospital and Faculty of Medicine, Heidelberg, Germany; Division of Neurology, Hospital de Clínicas de Porto Alegre, Brazil; Sorbonne Universite, Institut du Cerveau Paris Brain Institute-ICM, Inserm, CNRS, AP-HP, Inria Aramis project-team, Paris, France; Euro-ataxia, London, UK; Fondation de l’Ataxie Charlevoix-Saguenay, Montréal, QC, Canada; EURO-HSP, Paris, France; Deutsche Heredo Ataxie Gesellschaft (DHAG), Stuttgart, Germany; Department of Clinical Neurosciences, University of Cambridge, Cambridge, UK; Department of Neurology, Knappschaft Kliniken Recklinghausen, Germany; Molecular Medicine for Neurodegenerative and Neuromuscular Diseases Unit, IRCCS Stella Maris Foundation, Pisa, Italy; School of Medicine, Department of Neurology, Koc University, Istanbul, Turkey; Department of Neurology, Donders Institute for Brain, Cognition and Behavior, Radboud University Medical Center, Nijmegen, the Netherlands; Centre de Recherche Charles-Le Moyne, Sherbrooke University, Sherbrooke, QC, Canada; German Center for Neurodegenerative Diseases (DZNE), Tübingen, Germany; Division Translational Genomics of Neurodegenerative Diseases, Hertie-Institute for Clinical Brain Research and Center of Neurology, University of Tübingen, Tübingen, Germany

**Keywords:** hereditary spastic paraplegia, spastic ataxia, patient focus, patient-focused drug development, fit-for-purpose outcome, patient preference

## Abstract

**Background:** As therapeutic options emerge for hereditary spastic paraplegias (HSP), clinical trials require outcome measures that reflect disease aspects most important to patients. Patient priorities in HSP remain poorly defined. This study aimed to develop a regulatory-compliant framework of patient-prioritised health domains to evaluate treatment response in clinical trials.

**Methods:** Patient-reported data on health impacts were collected via two multinational, multilingual online surveys conducted sequentially, including 616 and 504 patients across the clinical and genetic spectrum of HSP. Using a staged approach, we examined prevalence, relevance, and severity, focusing on health impacts that were (i) common (ii) sensitive to disease progression, (iii) highly relevant to patients, and (iv) showed strong severity-relevance correlation. Patient representatives contributed centrally to study design and prioritisation.

**Findings:** Our patient-focused analysis yielded five highly prevalent and relevant core health domains: mobility, lower body function, autonomic dysregulation, pain, and psychosocial aspects. Ambulation and lower body function ranked highest across all disease stages. Among non-motor impacts, reduced ability to work, bladder incontinence, and fatigue were most relevant. In mild disease stages, reduced walking distance, reduced walking speed, and the urgency to empty the bladder were the most frequent and most relevant health impact.

**Interpretation:** This work provides the most comprehensive patient-reported and disease stage specific profiling of HSP health impacts to date. It lays the necessary groundwork for developing patient-focused outcome tools capable of capturing treatment effects in future trials.

## Introduction

Hereditary spastic paraplegias (HSP) and spastic ataxias (SPAX) represent a heterogeneous group of rare neurodegenerative disorders characterised by progressive lower limb spasticity and weakness.^1^ Clinical symptoms range from pure motor symptoms to complex syndromes with frequent affection of the cerebellar and other neuronal systems.^2,3^ The clinical and genetic heterogeneity, small patient cohorts, together with a slow disease progression make it particularly challenging to develop sensitive and patient-relevant outcome measures capable of detecting disease progression and treatment response in HSP trials.^4^ A scoping review in 2023 identified more than 80 distinct outcome measures across HSP studies, revealing substantial variability, inconsistent application, and the absence of consensus guidelines.^5^ While established clinician-reported outcome measures (ClinRO), such as the Spastic Paraplegia Rating Scale (SPRS), reliably detect disease change over time,^6^ they may not fully capture the breadth of symptoms, and the lived experiences that patients rate most meaningful. Regulatory bodies such as the FDA and EMA have called for a patient-focused approach to drug development and emphasise the need to integrate patient experiences and priorities into endpoint selection.^7^ Patient-reported outcome measures (PROM) thus have evolved as means to capture health impacts directly from the HSP community.^5,8,9^ However, fit-for-purpose outcome measures (whether ClinROs, PROMs, or performance-based assessments) must be designed to serve a specific context of use (e.g. a clinical trial) and reflect health impacts relevant to the targeted patient population. Still, representative ‘patient voice’ studies in HSPs, in particular those taking into account disease stage specific shifts in patient experiences and priorities, are missing. To address this gap and mismatch between what is measured and what matters to patients, we designed a large-scale concept elicitation study in collaboration with the PROSPAX consortium. We conducted two multinational, multilingual online surveys collecting anonymous patient-reported data on an exhaustive list of health impacts across the spectrum of HSP and ataxias. The present analysis, including over 600 participants, only focuses on patients with HSP and SPAX, which we considered within the broader HSP spectrum. The resulting dataset represents the most comprehensive patient-reported study in HSP/SPAX to date. Using a staged approach, we systematically explored the prevalence, relevance, and severity of a wide range of health impacts. Guided by the intended clinical trial context of use, our analysis prioritised domains that met the following criteria: they were (i) highly prevalent in the cohort, (ii) sensitive to disease progression, (iii) rated as highly relevant by patients, and (iv) demonstrated a strong correlation between severity and relevance. By capturing both the prevalence and prioritisation of the health impacts that were most meaningful to HSP patients, this study establishes a comprehensive, disease-stage specific, patient-focused framework that can be applied to guide the development and selection of fit-for-purpose outcome measures in upcoming clinical trials.

## Methods

### The PROSPAX patient-focused outcomes working group (PFOM working group)

PROSPAX (“PROSPAX – an integrated multimodal progression chart in spastic ataxias”; NCT04297891) is an international collaborative research project funded by the European Joint Programme on Rare Diseases (EJP-RD; grant agreement 441409627, 2020 – 2024). The PROSPAX PFOM working group is coordinated by Euro-ataxia (https://www.ataxia.org.uk) and consists of representatives from patient organisations (Euro-HSP, Euro ataxia, Ataxia UK, ARSACS Foundation, Deutsche Heredo-Ataxie Gesellschaft [DHAG]), PROSPAX patient partners (eight individuals with ataxia, SPAX and HSP), as well as principal investigators from the consortium. The study’s overall strategy, including the design of the surveys, analysis, and presentation of the survey results, was developed collaboratively by the PROSPAX PFOM working group.

### Multilanguage multi-national survey to collect health impacts of HSP (“Survey 1”)

The PROSPAX PFOM working group designed an anonymous survey to capture disease-associated symptoms and health impacts in individuals with HSP, SPAX and ataxia. The survey included items on demographics (sex, age), medical diagnosis (ataxia, HSP, other/free text), genotype, disease stage (ranging from no walking device usage to full-time wheelchair dependence) and health impacts. The initial list of 69 health impacts provided in the survey was compiled by combining suitable items from a published member survey by Ataxia UK^10^ and complemented by a targeted Pubmed literature review of cohort studies from the past ten years in ataxia/SPAX/HSP as well as consensus from the working group. Free text options allowed participants to report additional symptoms or health experiences. For participants under 16 years and/or those unable to self-complete, proxies (caregivers or family members) could respond on their behalf. Data were collected through a SurveyMonkey (www.surveymonkey.com) questionnaire, available to participants between December 2020 and January 2021.

### Multilanguage multi-national survey to determine patient-reported severity and relevance of health impacts in HSP (“Survey 2”)

We next designed a follow-up survey to assess severity and relevance of health impacts identified in Survey 1. In line with the intended context of use in a clinical trial setting, we reduced the number of items from 69 to 36 by removing items that had ≥ eight % missing data (nine items), were reported by < 30% of participants over the disease course (four items) or were judged by the PROSPAX PFOM working group as unspecific or unlikely to change with disease progression (ten items). Items reflecting related concepts were combined, further reducing the number by 12. Although ‘bowel incontinence’ was reported by 22 % of participants in Survey 1, the item was retained as the PROSPAX PFOM working group considered this symptom to be particularly relevant in everyday life (addition of 1 item). Based on patient representative feedback, three broad items were split into separate questions to improve specificity (e.g. bladder incontinence separated into bladder urgency and urine leakage). Participants rated symptom severity over the past two months using a five-point Likert scale ranging from “not present” to “very severe”. Relevance was assessed by how Relevance was assessed by how strongly a health impact affected participants’ daily physical or emotional life, using a five-point Likert scale ranging from “no effect” to “very severe effect”. Disease severity was assessed using an adapted version of the Friedreich Ataxia Rating Scale Functional Staging (FARS Staging)^11^ as a patient-reported measure (Roller et al. in preparation). Data were collected anonymously through the platform EUSurvey (https://ec.europa.eu/eusurvey/) between January – February 2022. Ataxia UK’s Internal Ethics Committee reviewed and approved the wording of both surveys prior to their distribution.

### Survey translation

Survey 1 and 2 were initially composed in English and translated into six languages (Canadian French, Dutch, French, German, Italian, and Turkish). Translations were performed by native speakers and independently reviewed by a second translator, with discrepancies resolved by the translation team. Free text responses from both surveys were translated into English for analysis by PFOM working group members. English versions of the surveys are provided in the supplement.

### Survey distribution

Surveys were distributed through the PROSPAX ataxia and HSP outpatient clinics and multiple patient organizations encouraging participation via their websites, social media, and newsletters. Participating organisations included ADCA Vereniging Nederland, Association Française Ataxie de Friedreich (AFAF), Associazione Italiana Sindromi Atassiche (A.I.S.A.), Associazione Italiana Vivere la Paraparesi Spastica (A.I.Vi.P.S.), Association Connaître les Syndromes Cérébelleux (CSC), Association Strümpell-Lorrain France, Ataxia Charlevoix-Saguenay Foundation, Ataxia UK, Deutsche Heredo-Ataxie Gesellschaft (DHAG), Euro-ataxia, Forum Ge(h)n mit HSP, Friedreich Ataxie Förderverein e.V., HSP Research Foundation, HSP Selbsthilfegruppe e.V., HSP UK support group, Spastic Paraplegia Foundation, Spierziekten Nederland, Tom Wahlig Foundation, and the Turkish Association of Muscle Diseases.

### Exclusion of responses

Only individuals reporting a diagnosis of HSP or SPAX were included in the analysis. Respondents reporting other diagnoses, including those with ataxia (n=491 in Survey 1, n=294 in Survey 2) or other neurological disorders (e.g. amyotrophic lateral sclerosis, multiple system atrophy, Charcot-Marie-Tooth disease), were removed. Participants missing mobility status data (n=4 in Survey 1, n=11 in Survey 2) that prevented disease stage classification were excluded from disease stage-specific analyses. In Survey 2, three participants with a genetic HSP diagnosis but no current mobility limitations were considered pre-symptomatic and excluded from further analyses. Of note, data on ataxia will be reported elsewhere.

### Disease classification of survey respondents

Hereditary ataxias and HSPs share considerable clinical and genetic overlap. Respondents were classified as predominant ataxia (ATX) or predominant HSP (HSP-predominant) based on reported genotypes (Table S1). SPAX represent an overlap phenotype within the ataxia-HSP spectrum; the subgroup was based on gene mutations linked to cerebellar and pyramidal features. For participants without a genetic diagnosis, classification was based on self-reported diagnosis.

### Grouping into disease stages

Respondents were classified into mild, intermediate and advanced stage based on overall mobility. Classification details are provided in the supplement (Table S2, Crnkovic V et al., in preparation).

## Statistical analysis

Statistical analyses were performed in Python (v3.12.7) using pandas (v2.2.2) and NumPy (v1.26.4) for data processing.^12,13^ Descriptive statistics summarised health impact prevalence (Survey 1), severity and relevance ratings (Survey 2). Differences across disease stages and between HSP-predominant and SPAX groups were measured using chi-square tests (SciPy v1.13.1), with p-values < 0.05 considered statistically significant.^14^ Multiple comparisons were corrected using the false discovery rate (FDR) method (statsmodel v0.14.2), with adjusted p-values (q-values) reported in tables. Data visualisation used Matplotlib (v3.9.2) and Seaborne (v0.13.2).^15,16^ Suitability for exploratory factor analysis (EFA) was confirmed by Bartlett’s test of sphericity (Survey 1: χ² = 12343.26, *p* <.0001, Survey 2: χ² = 5024.31, *p* <.0001) and the Kaiser-Meyer-Olkin measure (KMO = 0.89 in Survey 1, KMO = 0.83 in Survey 2). ^17,18^ Participants with >20% missing responses were excluded (Survey 1: n = 556, Survey 2: n = 481); remaining missing values were imputed using item-wise medians. For comparability, Survey 2 severity items (Likert scale, 0-4) were dichotomised to symptom presence (1-4) versus absence (0). EFA used MINRES extraction with Varimax rotation ^19^, retaining factors with eigenvalues >1 and scree plot inspection. Loadings ≥0.40 were considered meaningful (factor_analyzer library v0.5.1).^20^ Polychoric correlations between severity and relevance in Survey 2 were computed in R (v4.4.3) using the polychor () function from the polycor package (v0.8.1) with heatmaps generated via ggplot2 (v3.5.2) package.^21,22^

## Results

### Prioritisation framework for health impact selection

A multi-step analysis was applied using tailored criteria to prioritise health impacts relevant for capturing treatment response in HSP clinical trials. Rather than presenting an exhaustive catalogue, we aimed to generate a clinically useful and regulatory-relevant set of health impacts for future research and outcome measure development. Survey results were prioritised using four criteria: (i) prevalence, identifying health impacts that are sufficiently common within the target population; (ii) disease stage sensitivity, identifying health impacts that vary with disease progression and may capture longitudinal change and treatment response; (iii) patient-perceived relevance, reflecting what matters most to patients’ daily living; and (iv) the correlation between severity and functional relevance, highlighting impacts where treatment-induced reduction of severity would translate into reduced impact on daily living. The following section details the application of this framework in our study cohort.

### Demographics of Survey Respondents

*Survey 1:* A total of 1107 responses were collected. After applying inclusion and exclusion criteria, 616 individuals were classified as HSP, including 205 (33.3%) with a SPAX phenotype. Classification into HSP-predominant and SPAX subtypes relied on known genotypes (434, 70.5%, with the remainder categorised by self-reported diagnoses (182, 29.5%) (Fig. 1a and 1c). Key demographic features are summarised in Table 1. Overall, 47.6% of participants were male, the mean age was 49.3 ± 17.6 years. Most participants completed the survey in English (34.6%) or German (32.3%), with additional representation from several other languages. Based on mobility classifications (Table S2), 24.7% were in a mild (n=152), 62.2% in an intermediate (n=383) and 12.5% in an advanced disease stage (n=77) (Fig. 1a). Proxy reports identified 39 (6.3%) participants aged 1–15 years (9.3 ± 3.9 years). The most common genotypes were SPG4 (*SPAST*, 30.0%), SPG7 (*SPG7,* 16.6%), ARSACS (*SACS,* 11.2%), with 29.5% lacking molecular diagnoses (Fig. 1c and Table S1).

**Figure 1.**
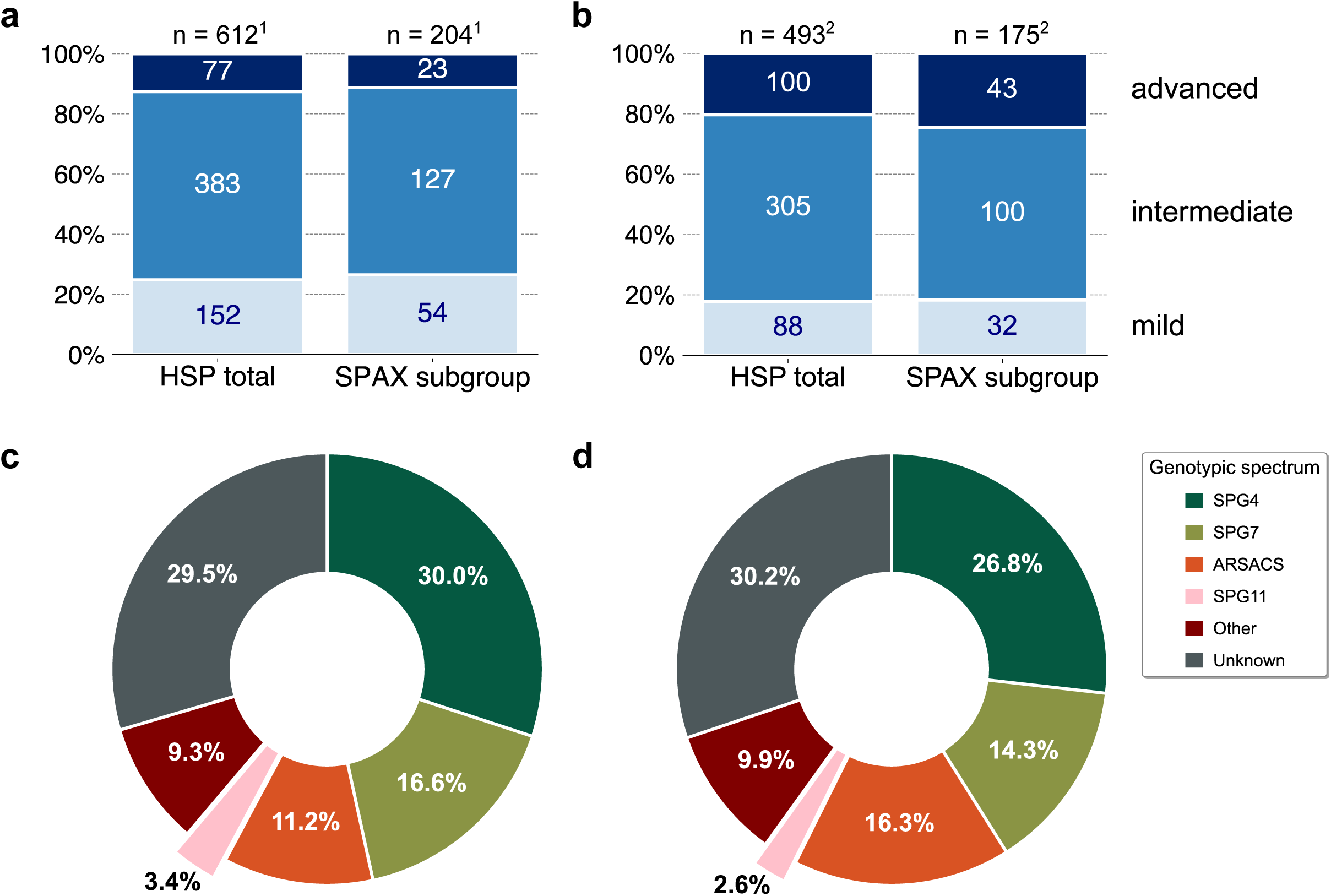
*Disease stages and genotypic spectrum.* **(a, b)** Stacked bar graphs illustrating the distribution of mobility-based disease stages (Table S2) for the overall cohort (left) and the SPAX subgroups (right) in Survey 1 **(a)** and Survey 2 **(b)**. Bar segments represent absolute numbers; bar heights reflect relative proportions (%).^1^ Missing disease stage data in Survey 1: HSP-predominant n=3, SPAX n=1.^2^ Missing disease stage data in Survey 2: HSP-predominant n=9, SPAX n=2. HSP, hereditary spastic paraplegia. SPAX, spastic ataxia. **(c, d)** Doughnut charts display genotype distribution as relative percentages in Survey 1 **(c)** and Survey 2 **(d)**. The “Other” category includes genetically defined HSP subtypes with n < 10 in the cohort (full list in Table S1). SPG, spastic paraplegia gene. ARSACS, autosomal recessive spastic ataxia of type Charlevoix-Saguenay.

**Table 1.** Demographic characteristics of survey respondents.

| Characteristic | Survey 1 |  |  | Survey 2 |  |  |
| --- | --- | --- | --- | --- | --- | --- |
|  | Adult | Children (proxy-reported) | Total cohort | Adult | Children (proxy-reported) | Total cohort |
| N | 577 (93.7 %) | 39 (6.3%) | 616 | 476 (94.4%) | 28 (5.6%) | 504 |
| Age (mean, standard deviation, range) | 52.1 ± 14.5 (16 - 82) | 9.3 ± 3.9 (1 - 15) | 49.3 ± 17.6 (1-82) | 52.4 ± 15.3 (16 - 85) | 9.2 ± 3.3 (5 - 15) | 50.0 ± 17.9 (5 - 85) |
| Sex (male / female / missing) | 269 (46.6%)<br>301 (52.2%)<br>7 (1.2%) | 24 (61.5%)<br>15 (38.5%)<br>0 (0.0%) | 293 (47.6%)<br>316 (51.3%)<br>7 (1.1%) | 220 (46.2%)<br>255 (53.6%)<br>1 (0.2%) | 15 (53.6%)<br>13 (46.4%)<br>0 (0.0%) | 235 (46.6%)<br>268 (53.2%)<br>1 (0.2%) |
| Disease stage (mild / intermediate / advanced / missing) | 132 (22.9%)<br>372 (64.5%)<br>70 (12.1%)<br>3 (0.5%) | 20 (51.3%)<br>11 (28.2%)<br>7 (17.9%)<br>1 (2.6%) | 152 (24.7%)<br>383 (62.2%)<br>77 (12.5%)<br>4 (0.6%) | 72 (15.1%)<br>297 (62.4%)<br>97 (20.4%)<br>10 (2.1%) | 16 (57.1%)<br>8 (28.6%)<br>3 (10.7%)<br>1 (3.6%) | 88 (17.5%)<br>305 (60.5%)<br>100 (19.8%)<br>11 (2.2%) |
| Language |  |  |  |  |  |  |
| *Canadian French* | 18 (3.1%) | 4 (10.3%) | 22 (3.6%) | 17 (3.6%) | 2 (7.1%) | 19 (3.8%) |
| *Dutch* | 42 (7.3%) | 0 (0.0%) | 42 (6.8%) | 56 (11.8%) | 2 (7.1%) | 58 (11.5%) |
| *English* | 195 (33.8%) | 18 (46.2%) | 213 (34.6%) | 194 (40.8%) | 17 (60.7%) | 211 (41.9%) |
| *French* | 76 (13.2%) | 2 (5.1%) | 78 (12.7%) | 72 (15.1%) | 1 (3.6%) | 73 (14.5%) |
| *German* | 195 (33.8%) | 4 (10.3%) | 199 (32.3%) | 112 (23.5%) | 1 (3.6%) | 113 (22.3%) |
| *Italian* | 44 (7.6%) | 10 (25.6%) | 54 (8.8%) | 16 (3.4%) | 4 (14.3%) | 20 (3.9%) |
| *Turkish* | 7 (1.2%) | 1 (2.6%) | 8 (1.3%) | 9 (1.9%) | 1 (3.6%) | 10 (2.0%) |

*Survey 2:* Overall, 801 individuals completed the second survey. After exclusions, 504 participants with HSP were retained for analysis, including 178 (35.3%) with SPAX. Respondents were classified into mild (17·5%, n=88), intermediate (60.5%, n=305) and advanced (19.8%, n=100) disease stages using FARS Staging (Fig. 1b, Table S2). Demographic characteristics are summarised in Table 1. Overall, 53.2% of respondents were male, the mean age was 50.0 ± 17.9 years. As in Survey 1, most participants completed the survey in English (41.9%) or German (22.3%), with smaller proportions responding in other languages. Proxy reports identified 28 participants aged ≤ 15 years (5.6% of the cohort). Self-reported genotype data revealed SPG4 (26.8%) as the most common subtype, followed by ARSACS (16.3%), and SPG7 (14.3%), with 30.2% of participants reporting unknown genotypes (Fig. 1d, Table S1).

### Prevalence of self-reported health impacts HSP

A total of 69 health impacts was assessed in Survey 1 using a self-administered questionnaire. Each item recorded whether an impact was currently present, previously experienced, or never experienced. To address criterion (i), prevalence of health impacts across the disease course was examined in the cohort (Fig. 2 and Table S3). On average, participants reported 31.8 ± 12.4 impacts. The ten most frequently reported impacts, listed in order of magnitude, were: reduced walking speed (96.9%), reliance on a handrail when ascending or descending stairs (95.7%), increased exertion while walking (94.5%), reduced walking distance (94.1%), balance issues while walking (94.0%), stumbling (93.8%), difficulty walking downhill (90.9%), leg weakness (89.0%), falling (86.7%) and muscle stiffness (85.2%). Overall, 42% of health impacts were reported by more than half of the cohort. These included core HSP-related features,^1^ such as impaired mobility, spasticity and urinary urgency, alongside broader, non-specific characteristics, such as fatigue (80.9%) and reduced ability to work (66.4%). In contrast, eight items were reported by fewer than 20% of participants (e.g. reduced visual field, 11.5%; epileptic seizures, 5.0%).

**Figure 2.**
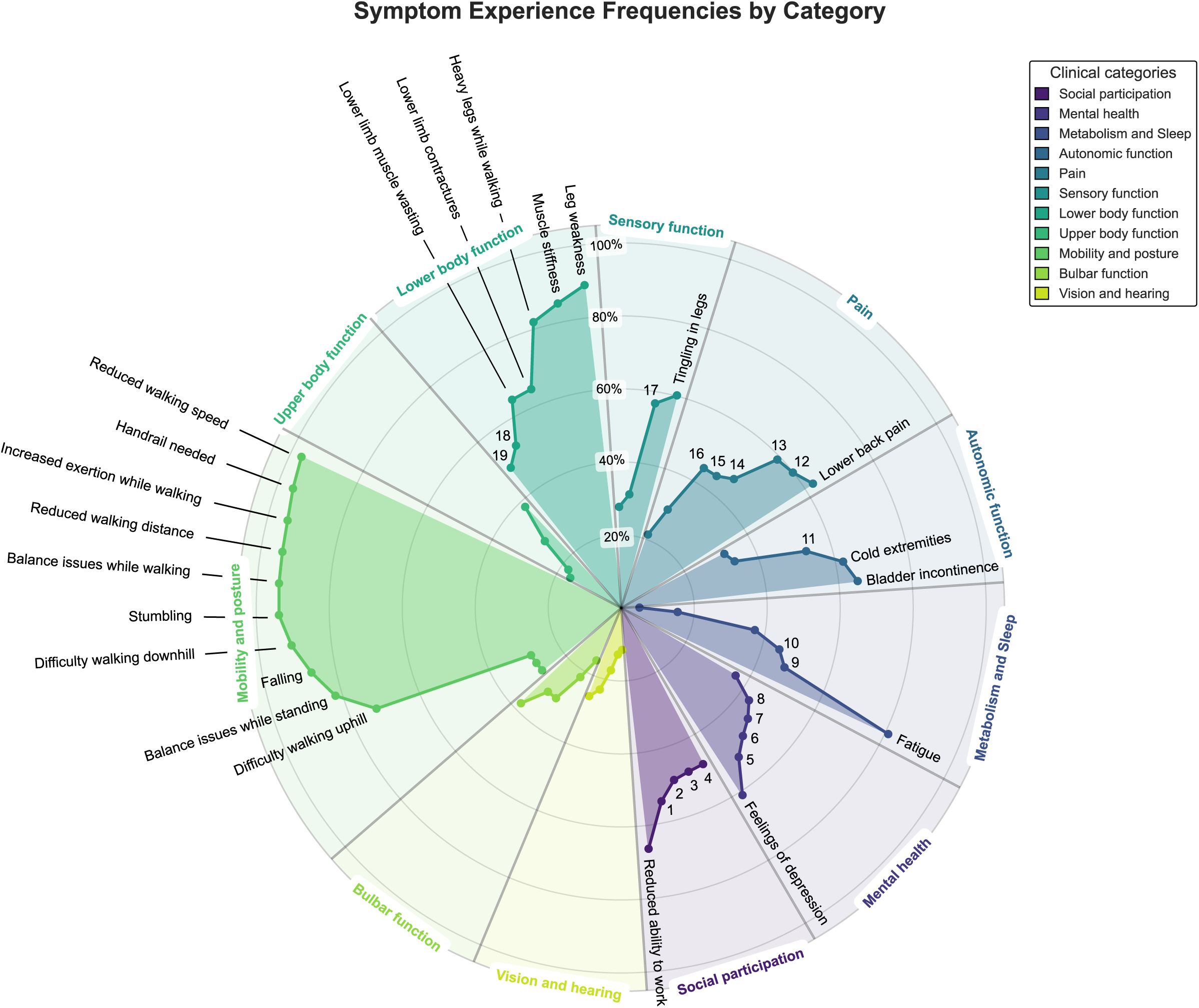
*Self-reported frequency of health impacts in HSP in Survey 1.* Spider plot with relative frequencies of health impacts experienced at any time over the disease course. Items are grouped by colour-coded functional domains (e.g. Mobility and posture). Frequencies are plotted as dots along a radial axis (0 – 100%). Commonly reported items (≥ 60%) are labelled by name; less frequent items (40 – 60%) are indicated by numbered markers: (1) Difficulty with social activities, (2) reduced sexual interest, difficulty meeting needs of (3) family and (4) friends, (5) lack of confidence, (6) poor concentration, (7) anxiety, (8) memory issues, trouble (9) staying and (10) falling asleep, (11) bladder emptying issues, (12) leg/foot pain, (13) pain while walking, (14) hip/groin pain, (15) neck/shoulder pain, (16) pain while standing, (17) numbness in legs, (18) muscle cramps and (19) claw toes. For cohort-wide comparability, the sex-specific health impact erectile dysfunction was excluded from the plot. A full list of all health impacts with corresponding counts and frequencies is provided in Table S3.

Health impacts were grouped into broader clinical domains. Items related to mobility and posture (100%) and lower body function (99.2%) were most frequently reported. Exploratory comparisons of item reporting between males and females identified sex-related differences (Table S4). Female respondents more frequently reported cold extremities, fatigue, insomnia, and pain-related symptoms. This pattern is consistent with epidemiological studies and suggests sex-related differences in symptom experience or reporting rather than HSP-specific effects.^23–25^ In contrast, slowing of speech and dysarthria (slowing of speech male/female 61.8/43.5%; dysarthria 62.7/44.0%) were more commonly reported by males. Although not statistically significant in subgroup analyses, this pattern was likely driven by the SPAX subcohort

### Mobility-related health impacts predominate in mild and intermediate disease stages

To explore how health impacts evolve with disease severity (criterion ii), participants were stratified into mild, intermediate and advanced disease stages (Table S3). In the mild stage, the most commonly reported health impacts were related to mobility and posture. These included reliance on a handrail when ascending or descending stairs (78.5%), reduced walking speed (76.8%), balance issues while walking (76.0%), leg weakness (72.1%), stumbling (69.8%), increased exertion while walking (69.2%) and difficulty walking downhill (68.9%) (Fig. S1a). In the intermediate stage, these mobility-related items remained highly prevalent and occurred at higher frequencies. Impacts related to autonomic dysregulation (bladder incontinence and retention, erectile dysfunction, cold extremities), sensory function and pain also emerged in this stage (Fig. S1b). In contrast, the advanced stage, defined by regular wheelchair use or dependency, showed a distinct health impact profile. Lower body related functions such as muscle stiffness (80.6%), contractures (73.1%), and muscle wasting (72.1%) remained frequently affected. The prevalence of autonomic dysregulation, sensory function and pain further increased. In addition, several symptom clusters related to bulbar function (speech and swallowing) and upper limb function emerged. An increased frequency of overweight was also reported (45.6%) (Fig. S1c).

Some aspects of psychosocial functioning were reported at similar frequencies across all disease stages. These included feelings of depression/sadness/hopelessness (38.6%), anxiety (31.7%), and anger (22.3%), suggesting that these symptoms emerge early in the disease course. Other aspects of cognitive or adaptive functioning - such as lack of attention/concentration, confidence dealing with unexpected events, and memory problems – became more frequent with advancing disease. In contrast, reduced social participation (e.g. ability to work, meeting needs of family/friends, interest in sexual activity), showed a marked increase in frequency from the mild to the intermediate stage, but then plateaued across the intermediate and advanced stages. Their incline thus seems to coincide with the loss of independent walking (defining the onset of the intermediate stage). Detailed stage-specific frequencies of health impacts are given in Table S3.

### HSP health impact clusters identified by exploratory factor analysis

Building on observed frequency patterns, we performed exploratory factor analysis (EFA) on currently experienced health impacts from Survey 1 across the total cohort. EFA with varimax rotation identified five distinct clusters, collectively explaining approximately 29% of the total variance (Fig. S2). Factor 1 (Ataxia) explained 8.23% of variance (eigenvalue = 5.679) and included items reflecting cerebellar impairment. Factor 2 (Pain & Sensory) accounted for 7.63% of variance (eigenvalue = 5.266) and comprised mainly pain and sensory symptoms. Factor 3 (Mobility) contributed 5.21% of variance (eigenvalue = 3.593) and included ambulation-related items. Factor 4 (Psychosocial Functioning) explained 4.71% of variance (eigenvalue = 3.25), included items related to emotional distress (e.g. depression) alongside social difficulties. Interestingly, reduced ability to work loaded with psychosocial functioning rather than mobility. Factor 5 (Autonomic Dysregulation) explained 3.01% of variance (eigenvalue = 2.083) and included bladder and bowel dysfunction-related impacts. Survey 1 was followed by a second survey designed to refine prioritisation of health impacts in HSP.

### Patient-defined priorities of health impacts in HSP

Survey 2 evaluated a refined set of 36 health impacts selected from Survey 1. Participants rated the severity and relevance of each health impact’ to daily life using five-point Likert scales, with relevance assessed only among participants who currently experienced a given impact. To prioritise impacts based on patient-perceived relevance (criterion iii), items were ranked by the proportion of affected participants rating them as at least moderately relevant (score ≥ 2) (Fig. 3 and Table S5). In total, 29 impacts (80.6%) were rated as at least moderately relevant by ≥50% of affected participants. The highest-ranked impacts included reduced walking speed (87.4%), reduced walking distance (87.3%), leg weakness (82.6%), balance while walking (81.2%), lower limb contractures (80.1%), impaired ability to work (79.3%), and urgency to empty the bladder (76.2%). In contrast, health impacts related to upper body function, sensory functions, speech and swallowing were among the lowest rated in terms of perceived relevance.

**Figure 3.**
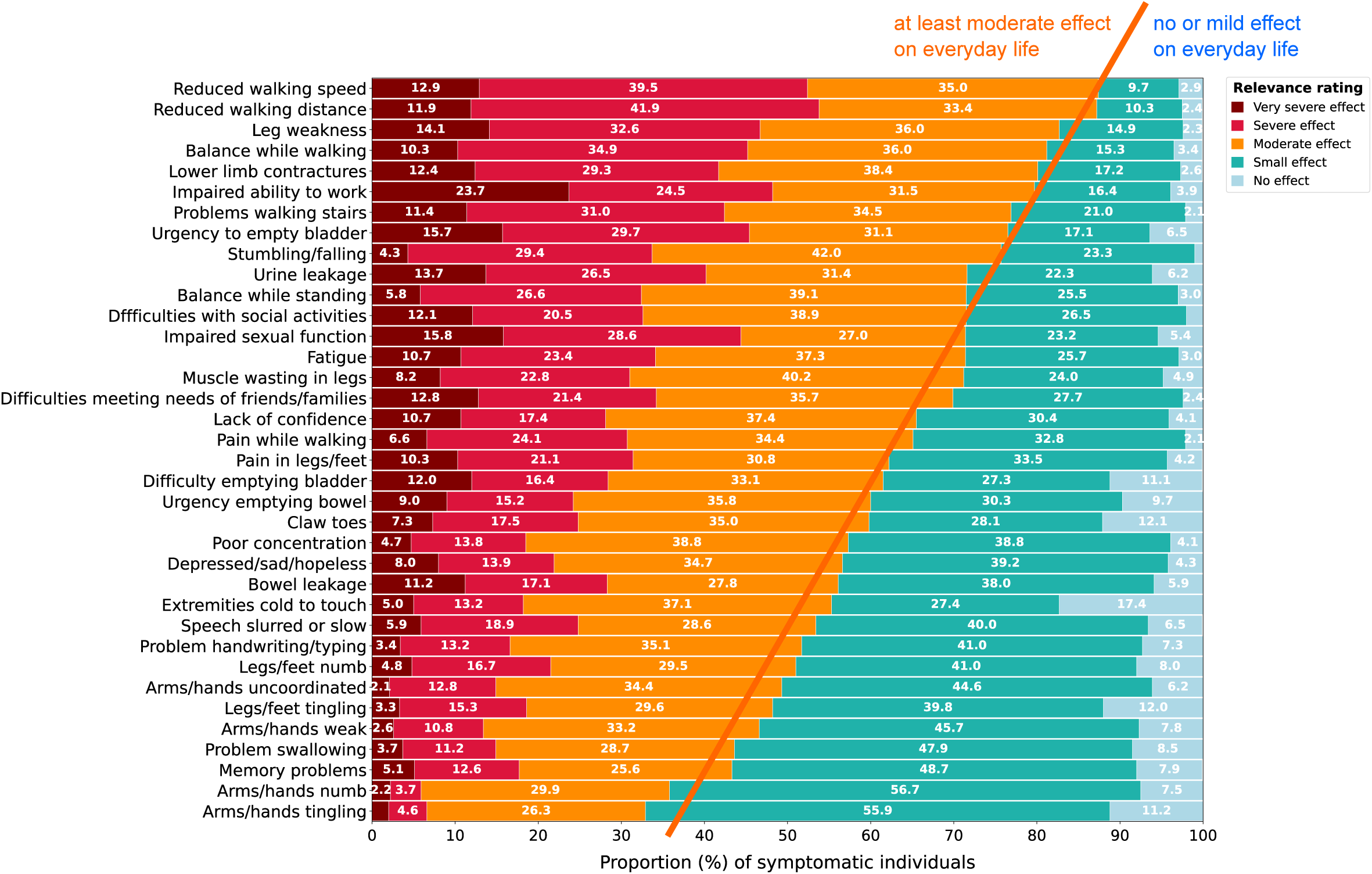
*Relevance of health impacts among symptomatic individuals in Survey 2.* Stacked bar chart illustrating the distribution of responses (%) related to symptom relevance only among those who experienced the symptom, ranging from “no effect” (light blue), “small effect” (turquoise), “moderate effect” (orange), “severe effect” (red), and “very severe effect” (maroon). Questions were ordered top-to-bottom by the combined percentage of moderate to very severe symptom relevance in a descending fashion.

Ambulatory individuals, representing the mild and intermediate disease stages, are likely candidates for future disease-modifying treatments. We therefore performed a subanalysis of perceived relevance within this ‘trial-relevant’ cohort (Fig. S3a). Ranking patterns largely mirrored that of the overall survey cohort: mobility and lower body function impacts remained top concerns, while impaired ability to work, fatigue and urgency to empty the bladder were also ranked highly. In comparison, individuals in advanced stages placed greater relevance on autonomic dysregulation, reduced social participation and other aspects of cognitive and adaptive functioning. These findings suggest that while prioritised health impacts are shared across the HSP population, their prevalence varies with disease stage. Functional loss in the later disease stages overlaps with an increased importance of psychosocial and emotional health aspects.

### Linking health impact severity to daily relevance in health impact prioritisation

To serve as meaningful outcome measures, health impacts must not only be common and relevant to patients but also show a clear link between experienced severity and perceived daily relevance (criterion iv). Only then can treatment-induced changes in severity translate into meaningful improvements in patients’ lives. To study this, we calculated polychoric correlations between severity (Fig. S4) and relevance ratings across all 36 impacts (Fig. S6, Table S6). Correlation coefficients ranged from 0.64 to 0.95 (median 0.86, IQR: 0.81 – 0.92), suggesting higher severity ratings generally coincided with greater perceived relevance. Particularly strong correlations (r > 0.9) were observed for impacts related to pain, upper body function, bulbar function and mental health. Slightly lower, but still robust correlations were seen for ambulation-related items, including reduced walking distance (r=0.75) and walking speed (r=0.77). As these mobility-related impacts did not apply to advanced-stage participants, correlations reflect the mild and intermediate subcohort only.

### Validation of health impact clusters in Survey 2 by exploratory factor analysis

To validate clusters identified in Survey 1, we performed exploratory factor analysis on the 36 prioritised health impacts rated in Survey 2. The analysis replicated a five-factor structure similar to Survey 1, explaining approximately 33.6% of total variance. Core domains, including Ataxia, Pain & Sensory, Mobility, Psychosocial Functioning, and Autonomic Dysfunction, were consistent with earlier findings and thereby demonstrated robustness of these clusters (Fig. S5).

*Prioritisation of core health domains for capturing meaningful treatment response in HSP* To prioritise health impact domains for capturing treatment response in HSP trials, we applied the above criteria: (i) prevalence, (ii) progression-related variation, (iii) patient relevance, and (iv) severity-relevance correlation. The integrated prioritisation is shown in a dot plot (Fig. 4), which maps symptom frequency on the x-axis and the percentage of participants experiencing at least moderate health impact on the y-axis. Dot sizes reflect the correlation between severity and relevance. Health impacts located in the upper-right quadrant are characterised by high frequency and high relevance. They represent candidate core impacts for detecting meaningful treatment change in clinical trials. Five core health domains were prioritised across the total cohort: mobility, lower body function, autonomic dysfunction, pain and social participation. Impacts particularly prominent in the mild disease stage (Fig. S7) were highlighted by red-circled markers to underline their potential suitability as early and sensitive outcomes. A 60% threshold for both frequency and relevance was applied to highlight impacts considered both common and meaningful to patients. This threshold was chosen pragmatically and can be adapted to specific study contexts. This approach results in a ranked yet flexible set of patient-focused core health domains in HSP. The adaptable nature of our framework accommodates varying contexts of use, including different study designs, disease stages, HSP subcohorts and future therapeutic targets.

**Figure 4.**
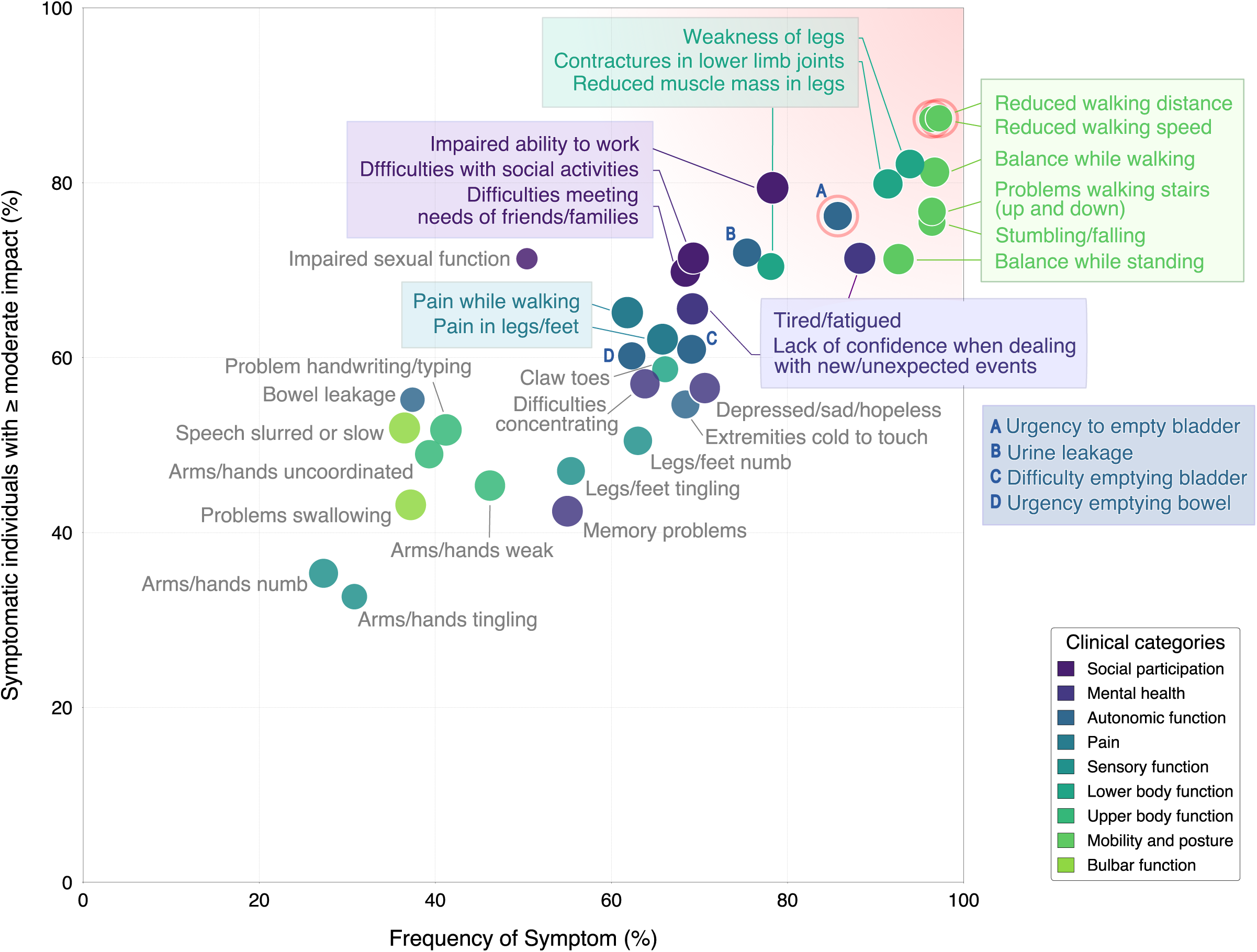
*Patient-prioritised core health impacts in HSP in Survey 2.* Scatterplot showing self-reported health impact frequency (x-axis) and the proportion of participants rating it as at least moderately relevant (y-axis). Dots are colour-coded by functional domains; dot size reflects the strength of correlation between health impact severity and relevance, Health impacts exceeding both 60% frequency and relevance are boxed and labelled; others are highlighted in grey. A gradient fade from the upper-right corner towards the origin represents decreasing health impact prevalence and relevance. Health impacts that are already highly prevalent and relevant in the mild disease stage are additionally marked with red circles, highlighting mild-phase targets. Disease stage specific-patterns are shown in *Fig. S6-S8*.

*Priority health impacts in ambulatory HSP patients (mild + intermediate disease stages)* In individuals with mild stage HSP, the most frequent and relevant impacts included reduced walking speed, reduced walking distance and bladder urgency (Fig. S7). With progression to intermediate stage, priority impacts expanded to a broader set of ambulation-related symptoms, including impaired balance while walking or standing, problems navigating stairs and stumbling/falling. Health impacts related to lower limb function, social participation and autonomic function also become more prevalent (Fig. S8).

### Priority health impacts in SPAX

Subanalysis of the SPAX cohort (Fig. S10) revealed a prioritisation profile broadly similar to that observed in the overall cohort. Mobility-related items remained the highest-priority domain, with reduced walking speed ranking among the most frequent and relevant symptom in individuals with mild-stage SPAX (data not shown). Items related to lower body function, autonomic dysregulation and cognitive and adaptive functioning were prioritised to a similar extent as in the total cohort. In contrast, pain-related items were less prominent and did not rank among the top domains. Although health impacts associated with cerebellar dysfunction were more than twofold more frequent in SPAX compared with the overall cohort (Table S3, modest shift of impacts to the right in Fig. S10), they did not reach the predefined prioritisation threshold. No other priority clusters were unique to SPAX. We thus concluded that prioritisation patterns closely mirrored those observed in the broader HSP cohort.

## Discussion

Through a patient-focused prioritisation framework tailored to support outcome selection in HSP/SPAX trials, we identified five core health domains that are most relevant to patients: mobility, lower body function, autonomic dysfunction, pain and social participation. We defined these areas as the most urgent unmet need in the field. While the raw data of this study may serve multiple future applications, our prioritisation of health impacts was guided by our context of use, that is, the prioritisation of health impacts for outcome selection in disease-modifying trials. Prioritised outcome domains in the SPAX subgroup closely resembled those in the total cohort.

This supports the idea of shared core outcome domains for HSP clinical trials despite phenotypic variation.

Importantly, this study was co-developed with patient representatives through the PFOM working group, whose input shaped the study design and prioritisation strategy. Patient relevance was embedded throughout the research process beyond survey responses, aligning with best practises for outcome development in rare disease research. As cultural background and socioeconomic context can influence how individuals perceive the impact of HSP on daily life, survey data were collected across HSP genotypes and cultural settings in collaboration with international partners. The resulting map of health impacts (Fig. 4) is applicable across HSP/SPAX subtypes and a large geographical region (Europe, North America, Australia) and remains adaptable to specific study needs.

For the majority of HSPs no disease-modifying therapies exist. Lack of accepted outcomes and endpoints contribute to this unmet need. Multiple candidate outcomes have been proposed, but none of these have yet demonstrated patient-relevance either for the concept itself or meaningful change. Specifically, a clear understanding of what matters to patients is needed in order to progress. Health impacts most relevant to HSP patients can then be used to select or develop meaningful outcomes, demonstrate the relevance of outcomes to patients, establish meaningful change with the goal of defining fit for purpose endpoints, align outcome measurement with both regulatory expectations and the priorities of the HSP community.

Our study therefore carries direct implications for outcome development in HSP. Outcome measures aiming to capture global disease burden can be anchored in health impacts highlighted in the upper right quadrant of Fig. 4, thereby ensuring that the resulting scale captures all relevant health impacts that are prevalent and meaningful across the phenotypic and disease-stage spectrum of HSPs. In trials targeting specific disease stages, the selected health impacts may be tailored to the disease stages addressed in the study or trial.

Mobility related functions, specifically reduced walking distance and reduced walking speed, are frequently impaired and prioritised by patients even in mild disease stages and remain the top-ranked health impacts throughout intermediate stages. Outcome measures specifically targeting these domains are therefore strong candidates for disease-modifying trials in mild (and intermediate) disease. It remains to be established, whether these concepts are best captured through digital mobility assessments, such as wearable sensors, or through alternative modalities, including performance outcomes. When analysing digital gait measures, these findings may direct the parameter space to be analysed.

Notably, patient-perceived balance impairments, typically reflecting deficits in coordination rather than pyramidal dysfunction, are highly relevant for HSP patients, supporting the use of patient-reported outcomes that capture balance confidence rather than solely objective balance performance.

Mobility-related items show ceiling effects in advanced disease stages due to functional loss and/or compensatory adaptations (e.g. wheelchair use). Consequently, autonomic dysfunction, lower limb impairments (e.g. claw toes, contractures), and reduced psychosocial functioning dominate the disease burden. These findings underscore that, beyond global measures of disease severity, advanced HSP requires stage-specific outcome measures, distinct from those used in earlier disease stages.

The most widely used clinical outcome assessment in HSP – the SPRS – primarily captures functional mobility, complemented by items addressing pain, contractures, and bladder function. As a global disease-severity scale, it is therefore well aligned with patient-prioritised health impacts. However, it does not adequately capture the impact of HSP on psychosocial participation and broader neurocognitive functioning. Newly developed outcome measures targeting disease-specific quality of life (TreatHSP-QoL) aim to address this gap, although their sensitivity to change remains to be established.^8^

Beyond selection of patient-focused outcome measures, the definition of trial endpoints requires the establishment of meaningful within-patient change. Consistent with regulatory guidance, anchor-based methods are central to deriving clinically meaningful change thresholds. This study is intended to guide and catalyse the development of patient-meaningful anchor measures capturing the most relevant health impacts in HSP. A consensus on such anchors will be essential to advance trial readiness in HSPs. The scope of highly relevant outcome domains hereby stressed the need for domain-specific anchor measures in addition to global anchors such as the Clinical Global Impression (CGI) or Patient Global Impression (PGI) tools.

Among non-motor health impacts, reduced ability to work, urinary incontinence, and fatigue were the most prevalent and relevant. Interestingly, impaired ability to work ranked highly in both overall and ambulatory (=mild + intermediate) cohorts (Fig. 3 and S3a). Factor analysis showed that this domain clustered with broader neurocognitive functions, suggesting that limitations are driven primarily by neurocognitive rather than mobility-related functions (Fig. S2 and S5).

Accordingly, improving psychosocial participation may require interventions targeting these domains, rather than approaches focused predominantly on mobility. Our findings also provide a framework for symptomatic treatment development, enabling clinicians and researchers to prioritise health domains that matter most to patients.

This study is not without limitations. While the data are representative for the group of HSP as a whole, individual priorities by genotypes, age or on the individual level may differ and our geographic catch is mostly representative of Europe, North America and Australia. Nevertheless, our workflow is inclusive and gives guidance for individual evaluation of relevance by context of use. Group classification relied on self-report of genotypes, age and disease type, which could affect accuracy. Furthermore, clinical importance of certain health impacts, such as impaired sexual function, may not be adequately captured by their frequency or severity ratings in survey data. While they are less commonly reported within the total cohort, if present, they are rated as highly relevant to their daily life (Table S5).

In summary, our results represent the most comprehensive assessment of patient-reported health experiences in HSP to date. Our work directly informs selection and development of meaningful outcomes and endpoints for disease-modifying trials across all stages of HSP. Moreover, the resulting framework and dataset offer a foundation for key stakeholders in the HSP field to better understand and prioritise what matters most to individuals living with HSP, supporting a wide range of contexts and applications.

## Contributors

M.A. and C.DF. accessed and verified the underlying study data and contributed to the initial design and execution of the formal analysis, supported by S.TdM. and R.S. Data curation was performed by S.TdM. and C.DF. M.A. led the data analysis strategy, was responsible for data visualisation, and prepared the original and final drafts of the manuscript together with R.S.. C.D., R.W., C.G., L.RL., A.N., R.H., S.K., F.M.S., A.V., B.vdW., C.G., J.MS., M.S., S.TdM., R.S. and the PROSPAX PFOM working group conceptualised and executed the study. J.MS., M.S., S.TdM. and R.S. also contributed to the organisation of the study. All authors performed critical review of the manuscript and approved the final version.

## Data sharing statement

The data used can be made available upon reasonable request and approval from the corresponding author.

## Declaration of interests

This work was supported by the Bundesministerium für Forschung, Technologie und Raumfahrt (BMFTR) through funding for the TreatHSP network (No. 01GM2209; to R.S. and S.K.), by the Deutsche Forschungsgemeinschaft (DFG) under the frame of the European Joint Programme on Rare Diseases (EJP RD) for the PROSPAX consortium (No. 441409627; to M.S., R.S., F.M.S., C.G., A.V., B.vdW., S.TdM.), the European Health and Digital Executive Agency (HADEA) through funding for the European Rare Disease Research Alliance (ERDERA) (grant agreement 101156595; to R.S. and M.S.), the EJP RD COFUND-EJP No. 825575 for the innovation project EVIDENCE RND (No. 825575 to M.S. and R.S.) and the Else Kröner Fresenius Stiftung through funding for the Clinician Scientist Programme PRECISE.net (M.S. and R.S.). F.M.S. is partially supported by the Italian Ministry of Health, Ricerca Corrente 2025-2026 and Telethon grant GJC21131. Several authors (M.S., R.S.) are members the European Reference Network for Rare Neurological Diseases – Project ID 739510.

## Data Availability

The data used can be made available upon reasonable request and approval from the corresponding authors.

## Acknowledgements

We are grateful to the PROSPAX patient partners for their active and invaluable participation in the PROSPAX PFOM working group. In particular we acknowledge Phil Jones, Lori Renna Linton, Karin Chladek, Carol McCudden, Caroline Pernon, Henry Wahlig, Claudia Maltais, and Carlo Carbotti. Moreover, we like to thank volunteers involved in translating both surveys: Ilse Willemse, Jorik Nonnekens (Dutch), Nazlı Başak, Ayca Sahin, Nazan Akkaya (Turkish), Filippo Santorelli, Susanna De Luca, Ivana Ricca (Italian), Chantal Gobeil, Valérie Gagné-Ouellet (Canadian French), Elodie Petit, Hortense Hurmic, Anas El Benna, Sophie Tezenas Du Montcel (French), Matthis Synofzik, Rebecca Schüle, Christoph Kessler, Andreas Nadke (German). Lastly, we are grateful to all patient organisations that distributed the survey among their members (see methods section for a list of collaborating patient organisations).

## Financial Disclosures

M.A., F.M.S., C.DF, C.G., S.K. and R.S report no disclosures. BvdW reports research support from ZonMw, FARA, Dutch Scientific Organization, Christina Foundation, Hersenstichting, and Servier; consultancy for or role in scientific advisory board for VICO Therapeutics and Biogen; and royalties from BSL/Springer Nature. R.H. is supported by the Wellcome Discovery Award (226653/Z/22/Z), the Medical Research Council (UK) (MR/V009346/1), the Hereditary Neuropathy Foundation, the AFM-Telethon, the Ataxia UK, the Action for AT, the Muscular Dystrophy UK, the Rosetrees Trust (PGL23/100048), the LifeArc Centre to Treat Mitochondrial Diseases (LAC-TreatMito) and the UKRI/Horizon Europe Guarantee MSCA Doctoral Network Programme.

## Appendix A. Supplementary data

**Supplementary Table 1.** Genotypes of survey respondents in Survey 1 and 2.

**Supplementary Table 2.** Disease stage classification in Survey 1 and 2.

**Supplementary Table 3.** Prevalence of self-reported health impacts in HSP, grouped by category [Survey 1].

**Supplementary Table 4.** Prevalence of self-reported health impacts in HSP, grouped by gender [Survey 1].

**Supplementary Table 5.** Severity and relevance of health-related impacts in daily life.

**Supplementary Table 6.** Polychoric correlation between severity and relevance.

**Supplementary Figure 1.** *Health impact frequency across disease stages in HSP [Survey 1].* Only those with complete mobility data (n=612, 99.4%) were included in stage-specific analyses. **(a, b, c)** Spider plots with relative frequencies of health impacts experienced at the time of the survey across mild **(a)**, intermediate **(b)**, and advanced **(c)** disease stages. Items are organised by functional domains and consistently ordered based on frequency in the mild stage (a) to enable comparison across plots. Frequencies are plotted as dots along radial axes (0–100%).

**Supplementary Figure 2.** *Heatmap of standardised factor loadings from exploratory factor analysis (EFA) of Survey 1 data (n = 556, varimax rotation).* Rows represent individual items; columns represent the extracted factors. The colour gradient reflects the strength of association (factor loading) between each item and factor. Factors were labelled based on thematic clustering of high-loading items: Ataxia (Factor 1), Pain & Sensory (Factor 2), Mobility (Factor 3), Psychosocial Functioning (Factor 4), and Autonomic Dysfunction (Factor 5).

**Supplementary Figure 3.** Relevance of health impacts among (a) ambulatory (mild and intermediate disease stage) and (b) advanced stage HSP in Survey 2. Layout and convention as in Fig. 4.

**Supplementary Figure 4.** *Distribution of self-reported symptom severity in Survey 2.* Stacked bar chart illustrating the distribution of responses (%) related to symptom severity, ranging from “symptom not present” (grey), “mild” (sand), “moderate” (mint green), “severe” (leaf green), and “very severe” (forest green). Questions were ordered top-to-bottom by the combined percentage of moderate to very severe symptom severity in a descending fashion.

**Supplementary Figure 5.** *Heatmap of standardised factor loadings from exploratory factor analysis (EFA) of Survey 2 data (n = 481, varimax rotation).* Rows correspond to individual health items; columns represent extracted latent factors. The colour gradient reflects the strength of association between items and factors. Factors labelled were based on thematic grouping of items with high loading: SPAX (Factor 1), Pain & Sensory (Factor 2), Mobility (Factor 3), Psychosocial Functioning (Factor 4), and autonomic dysfunction (Factor 5).

**Supplementary Figure 6.** Heatmaps of Likert-scale responses conveying participants’ ratings of severity versus relevance for health impacts in Survey 2, grouped by clinical category. Participants rated severity for all items; relevance ratings were provided only when the symptom was present, severity ≥1). Cells represent the percentage of total responses for each severity–relevance combination.

**Supplementary Figure 7.** Patient-prioritised core health impacts in mild stage HSP. Scatterplot layout and conventions as in Fig. 5.

**Supplementary Figure 8.** Patient-prioritised core health impacts in intermediate stage HSP. Scatterplot layout and conventions as in Fig. 5.

***Supplementary Figure* 9.** *Patient-prioritised core health impacts in advanced stage HSP.* Scatterplot layout and convention as in Fig. 5. A green box marks ambulation-related health items that cluster at the 0/0 origin, indicating that these are no longer reported as present or relevant by affected individuals.

**Supplementary Figure 10.** Patient-prioritised core health impacts in the SPAX subcohort. Scatterplot layout and conventions as in Fig. 5.

## Declaration of AI-assisted technologies in the manuscript preparation process

During the preparation of this manuscript the author used ChatGPT 4o in order to refine language of selected sentences. After using this tool, the author reviewed and edited the content as needed and takes full responsibility for the content of the published article.

## Notes

### Competing Interest Statement

This work was supported by the Bundesministerium fuer Forschung, Technologie und Raumfahrt (BMFTR) through funding for the TreatHSP network (No. 01GM2209; to R.S. and S.K.), by the Deutsche Forschungsgemeinschaft (DFG) under the frame of the European Joint Programme on Rare Diseases (EJP RD) for the PROSPAX consortium (No. 441409627; to M.S., R.S., F.M.S., C.G., A.V., B.vdW., S.TdM.), the European Health and Digital Executive Agency (HADEA) through funding for the European Rare Disease Research Alliance (ERDERA) (grant agreement 101156595; to R.S. and M.S.), the EJP RD COFUND-EJP No. 825575 for the innovation project EVIDENCE RND (No. 825575 to M.S. and R.S.) and the Else Kroener Fresenius Stiftung through funding for the Clinician Scientist Programme PRECISE.net (M.S. and R.S.). F.M.S. is partially supported by the Italian Ministry of Health, Ricerca Corrente 2025-2026 and Telethon grant GJC21131. Several authors (M.S., R.S.) are members the European Reference Network for Rare Neurological Diseases (Project ID 739510).

### Funding Statement

This work was funded by the Bundesministerium fuer Forschung, Technologie und Raumfahrt (BMFTR), Deutsche Forschungsgemeinschaft (DFG), European Joint Programme on Rare Diseases (EJP RD), European Rare Disease Research Alliance (ERDERA), and the Else Kroener Fresenius Stiftung (EKFS).

### Author Declarations

Ataxia UK's Internal Ethics Committee reviewed and approved the wording of both surveys prior to their distribution.

