## Supplementary Figures for "Identifying trial-relevant concepts of interest in HSP: insights from an international patient-voice study in over 600 individuals"

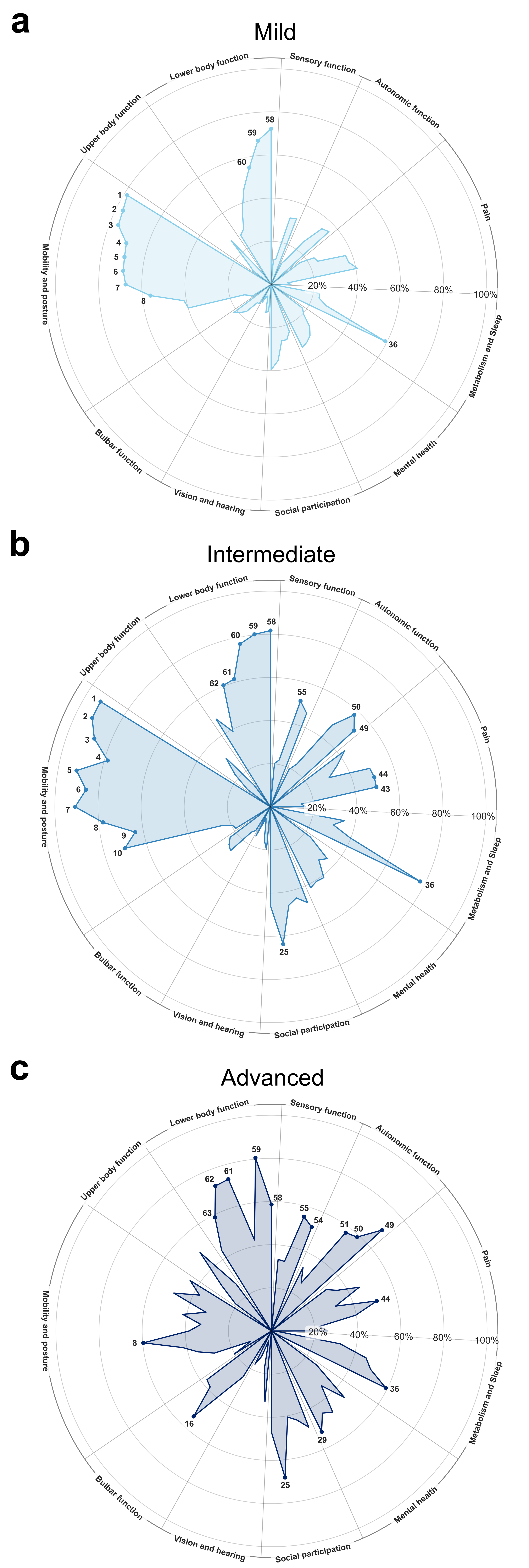


Supplementary Figure 1

| | Ataxia | Pain & Sensory | Mobility | Psychosocial Functioning | Autonomic Dysfunction |
|---|---|---|---|---|---|
| Reduced walking distance | -0.03 | 0.12 | 0.56 | 0.13 | 0.03 |
| Increased exertion while walking | 0.02 | 0.11 | 0.58 | 0.08 | 0.01 |
| Reduced walking speed | 0.01 | 0.05 | 0.58 | 0.06 | 0.09 |
| Stumbling | 0.08 | 0.04 | 0.38 | 0.01 | 0.07 |
| Falling | 0.07 | 0.06 | 0.38 | 0.05 | 0.06 |
| Pain associated with walking | 0.03 | 0.61 | 0.22 | 0.11 | 0.03 |
| “Heavy legs“ while walking | -0.05 | 0.42 | 0.31 | 0.09 | 0.01 |
| Weakness of legs | 0.02 | 0.26 | 0.41 | 0.05 | -0.08 |
| Problems with balance while walking | 0.05 | 0.05 | 0.46 | 0.08 | -0.00 |
| Dizziness while walking | 0.36 | 0.36 | 0.10 | 0.12 | 0.17 |
| Difficulty walking uphill | 0.08 | 0.24 | 0.39 | 0.13 | 0.11 |
| Difficulty walking downhill | 0.03 | 0.03 | 0.49 | 0.02 | 0.07 |
| Handrail needed when ascending or descending stairs | 0.05 | 0.04 | 0.48 | 0.08 | 0.13 |
| Problems with balance while standing | 0.18 | 0.11 | 0.41 | 0.11 | 0.07 |
| Dizziness while standing | 0.38 | 0.37 | 0.07 | 0.09 | 0.11 |
| Pain associated with standing | 0.08 | 0.55 | 0.15 | 0.04 | 0.12 |
| Weakness of arms | 0.65 | 0.27 | 0.14 | 0.01 | -0.03 |
| Clumsiness in arms/hands | 0.66 | 0.06 | 0.19 | 0.07 | -0.02 |
| Fatigue | 0.19 | 0.21 | 0.24 | 0.31 | 0.05 |
| Poor concentration | 0.35 | 0.19 | 0.07 | 0.33 | 0.13 |
| Feeling depressed/sad/hopeless | 0.12 | 0.24 | 0.10 | 0.56 | 0.13 |
| Regular feelings of anxiety | 0.24 | 0.30 | 0.06 | 0.55 | -0.02 |
| Regular feelings of anger | 0.23 | 0.20 | 0.06 | 0.46 | 0.05 |
| Oscillopsies | 0.49 | 0.16 | -0.02 | 0.11 | 0.24 |
| Double vision | 0.44 | 0.12 | -0.02 | 0.09 | 0.28 |
| Reduced visual field | 0.43 | 0.15 | -0.06 | 0.05 | 0.27 |
| Reduced visual acuity | 0.40 | 0.16 | -0.05 | 0.17 | 0.26 |
| Problems with swallowing | 0.48 | 0.13 | 0.13 | 0.25 | 0.01 |
| Drooling | 0.51 | 0.05 | 0.09 | 0.19 | 0.05 |
| Unable to speak loudly enough | 0.48 | -0.12 | 0.06 | 0.18 | -0.05 |
| Slurred speech | 0.66 | -0.15 | 0.10 | 0.19 | -0.06 |
| Slow speech | 0.66 | -0.14 | 0.15 | 0.19 | -0.13 |
| Numbness in arms/hands | 0.49 | 0.33 | 0.03 | 0.12 | -0.04 |
| Numbness in legs/feet/toes | 0.16 | 0.47 | 0.12 | 0.15 | 0.07 |
| Tingling in arms/hands | 0.46 | 0.42 | 0.00 | 0.11 | -0.02 |
| Tingling in legs/feet/toes | 0.07 | 0.56 | 0.13 | 0.17 | -0.01 |
| Frequent headache | 0.31 | 0.35 | 0.01 | 0.22 | 0.14 |
| Lower back pain | 0.05 | 0.48 | 0.12 | 0.14 | 0.16 |
| Neck/shoulder pain | 0.25 | 0.42 | 0.07 | 0.14 | 0.21 |
| Arm/hand pain | 0.45 | 0.46 | 0.01 | 0.10 | 0.05 |
| Hip/groin pain | 0.05 | 0.51 | 0.14 | 0.09 | 0.25 |
| Leg/foot pain | 0.05 | 0.66 | 0.16 | 0.13 | 0.04 |
| Upper limb contractures | 0.40 | 0.26 | 0.09 | 0.00 | 0.09 |
| Upper limb muscle wasting | 0.42 | 0.15 | 0.15 | 0.00 | 0.06 |
| Lower limb muscle wasting | 0.08 | 0.20 | 0.35 | 0.04 | 0.25 |
| Bladder incontinence | 0.06 | 0.07 | 0.17 | 0.04 | 0.50 |
| Difficulty emptying the bladder | 0.10 | 0.21 | 0.13 | 0.04 | 0.41 |
| Bowel incontinence | 0.10 | 0.12 | 0.11 | 0.04 | 0.44 |
| Constipation | 0.19 | 0.18 | 0.10 | 0.22 | 0.31 |
| Reduced sexual interest | -0.02 | 0.15 | 0.06 | 0.21 | 0.36 |
| Muscle stiffness | 0.11 | 0.31 | 0.26 | 0.15 | -0.04 |
| Muscle cramps | 0.22 | 0.36 | 0.19 | 0.15 | 0.01 |
| Overweight | -0.01 | 0.32 | -0.02 | 0.25 | 0.11 |
| Trouble falling asleep | 0.12 | 0.42 | 0.02 | 0.32 | 0.19 |
| Trouble staying asleep | 0.12 | 0.40 | 0.01 | 0.28 | 0.22 |
| Difficulty with social activities | 0.18 | 0.07 | 0.13 | 0.54 | 0.13 |
| Reduced ability to work | 0.13 | 0.15 | 0.31 | 0.40 | 0.14 |
| Difficulty meeting needs of friends | 0.16 | 0.13 | 0.23 | 0.45 | 0.13 |
| Difficulty meeting needs of family | 0.13 | 0.14 | 0.21 | 0.48 | 0.15 |
| Lack of confidence | 0.20 | 0.13 | 0.10 | 0.52 | 0.05 |

Supplementary Figure 2

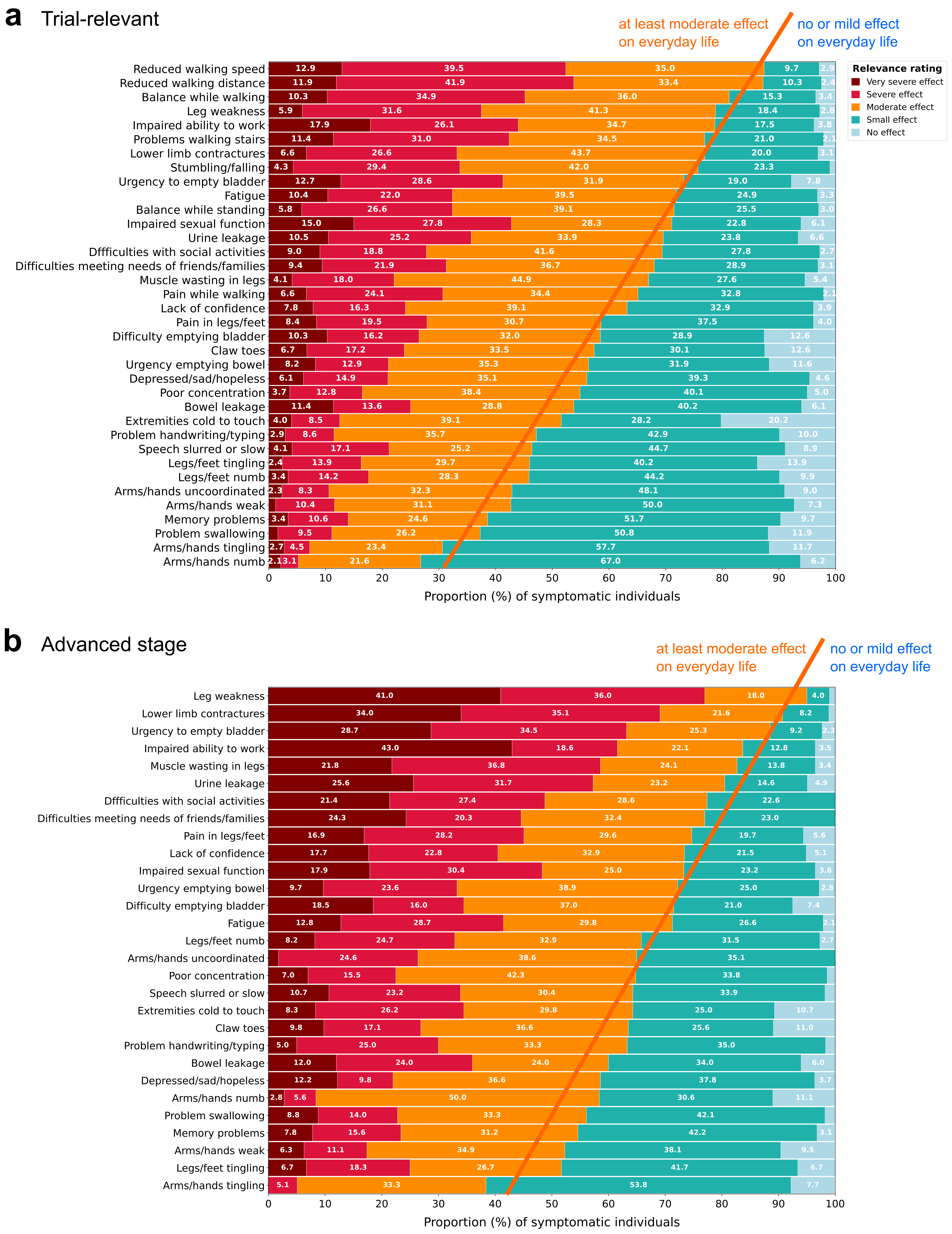


Supplementary Figure 3

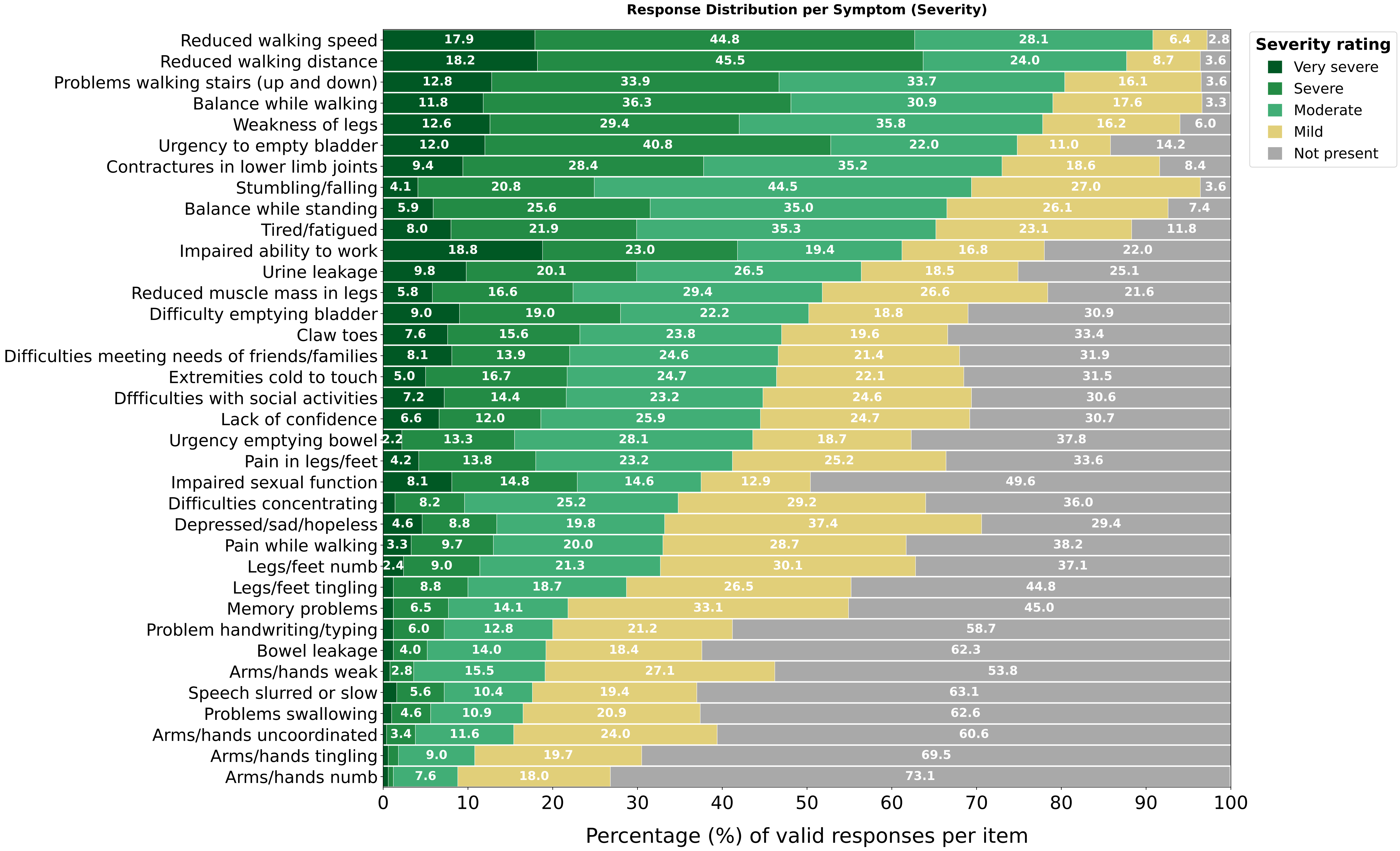


### Supplementary Figure 4

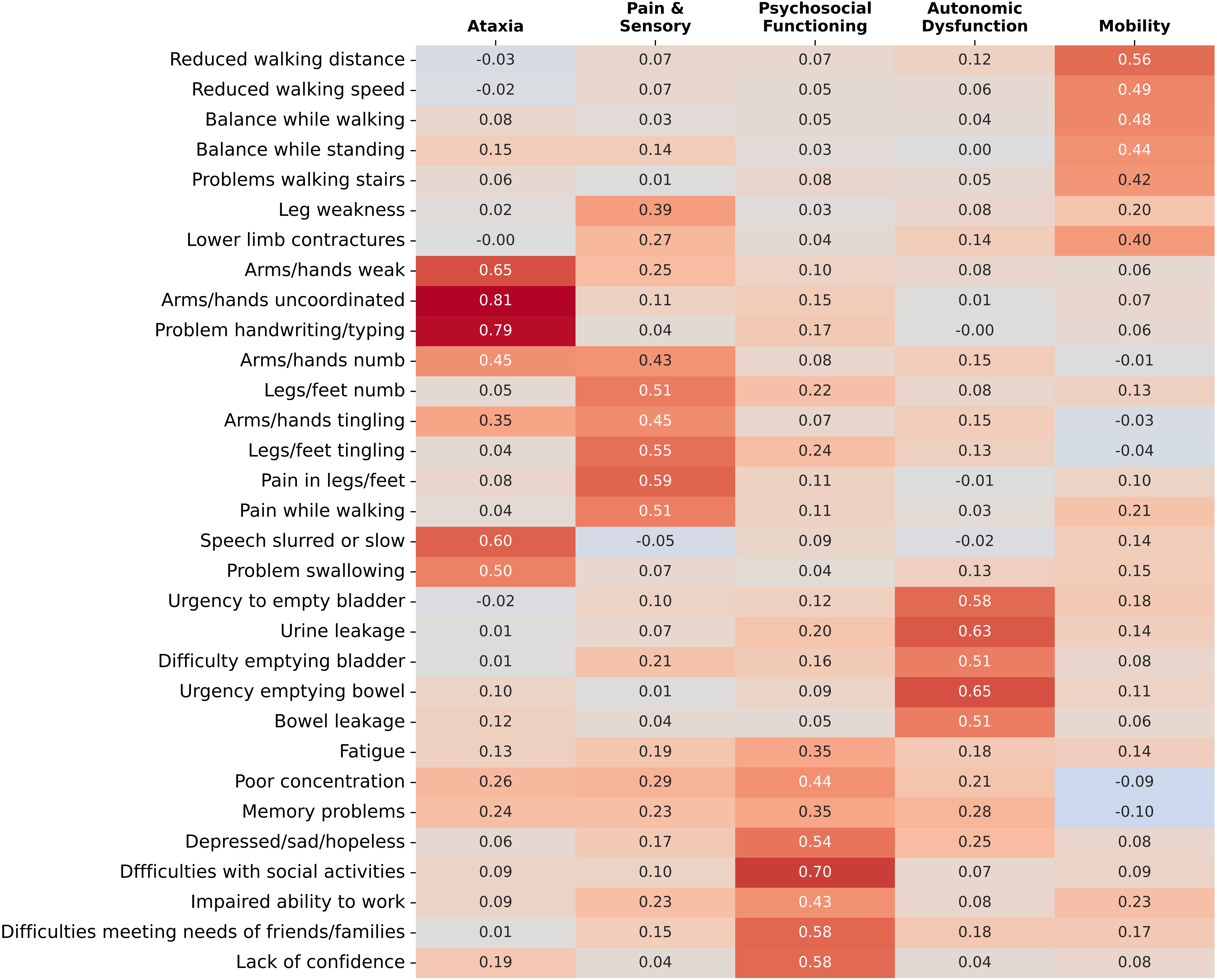


Supplementary Figure 5

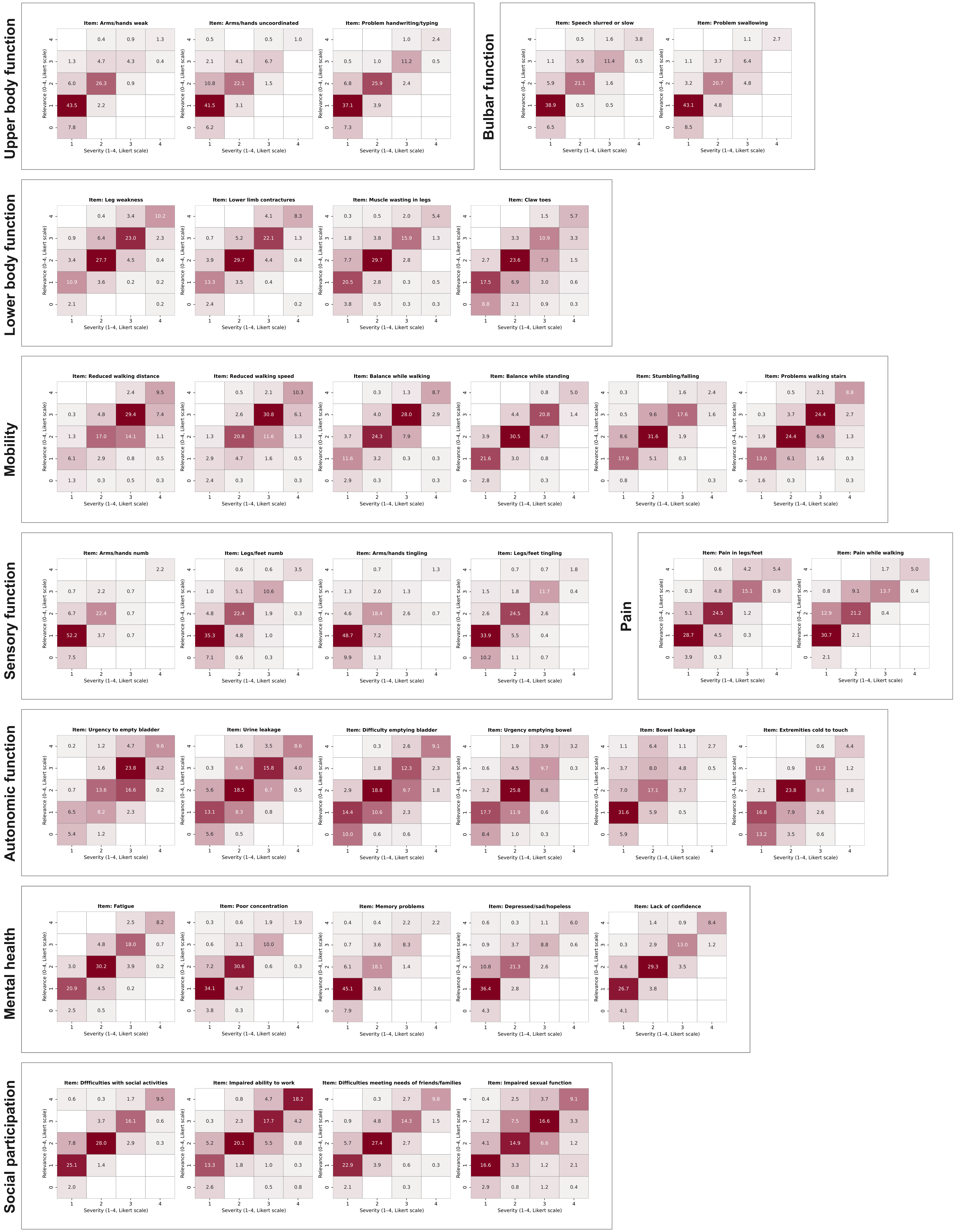


Supplementary Figure 6

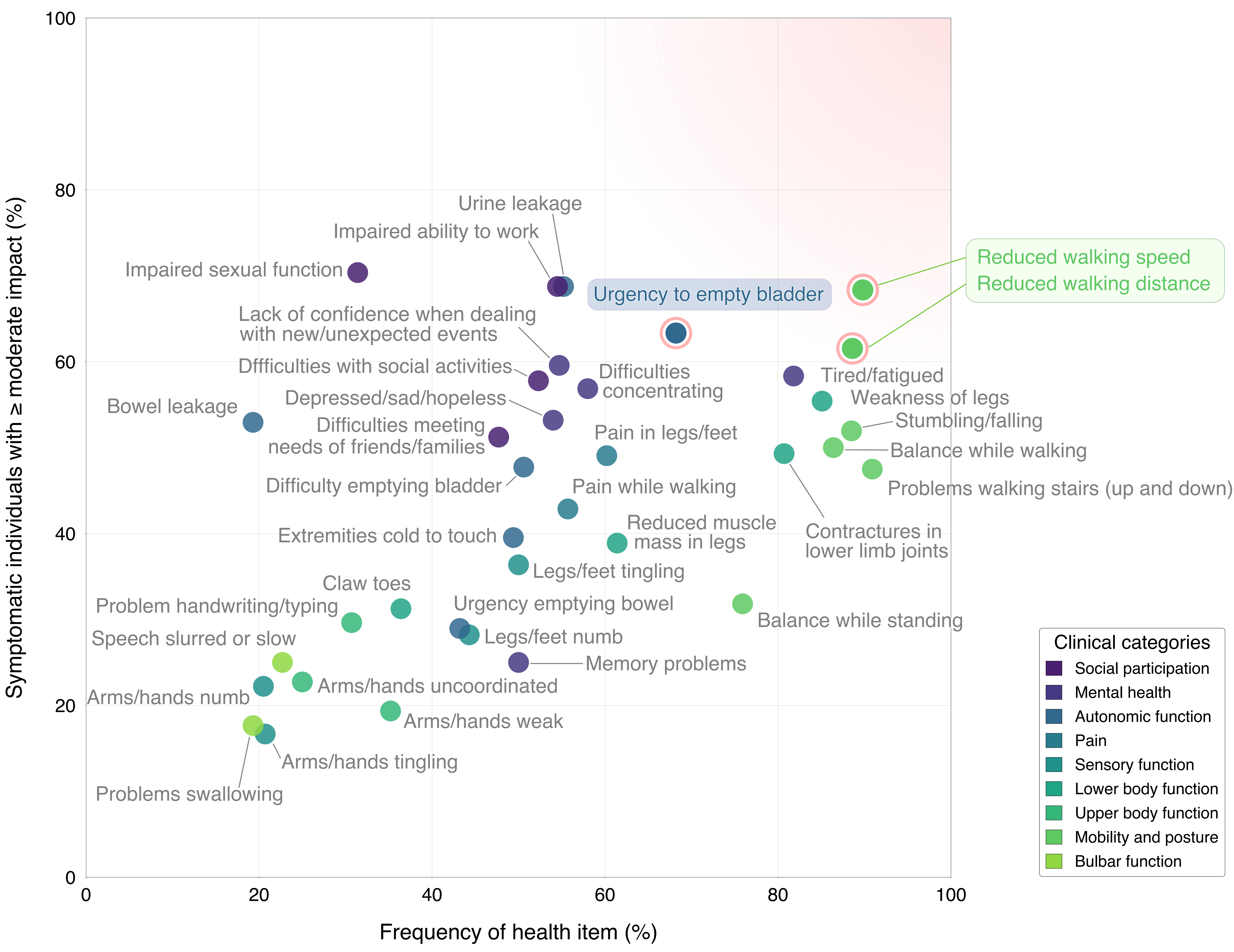


### Supplementary Figure 7

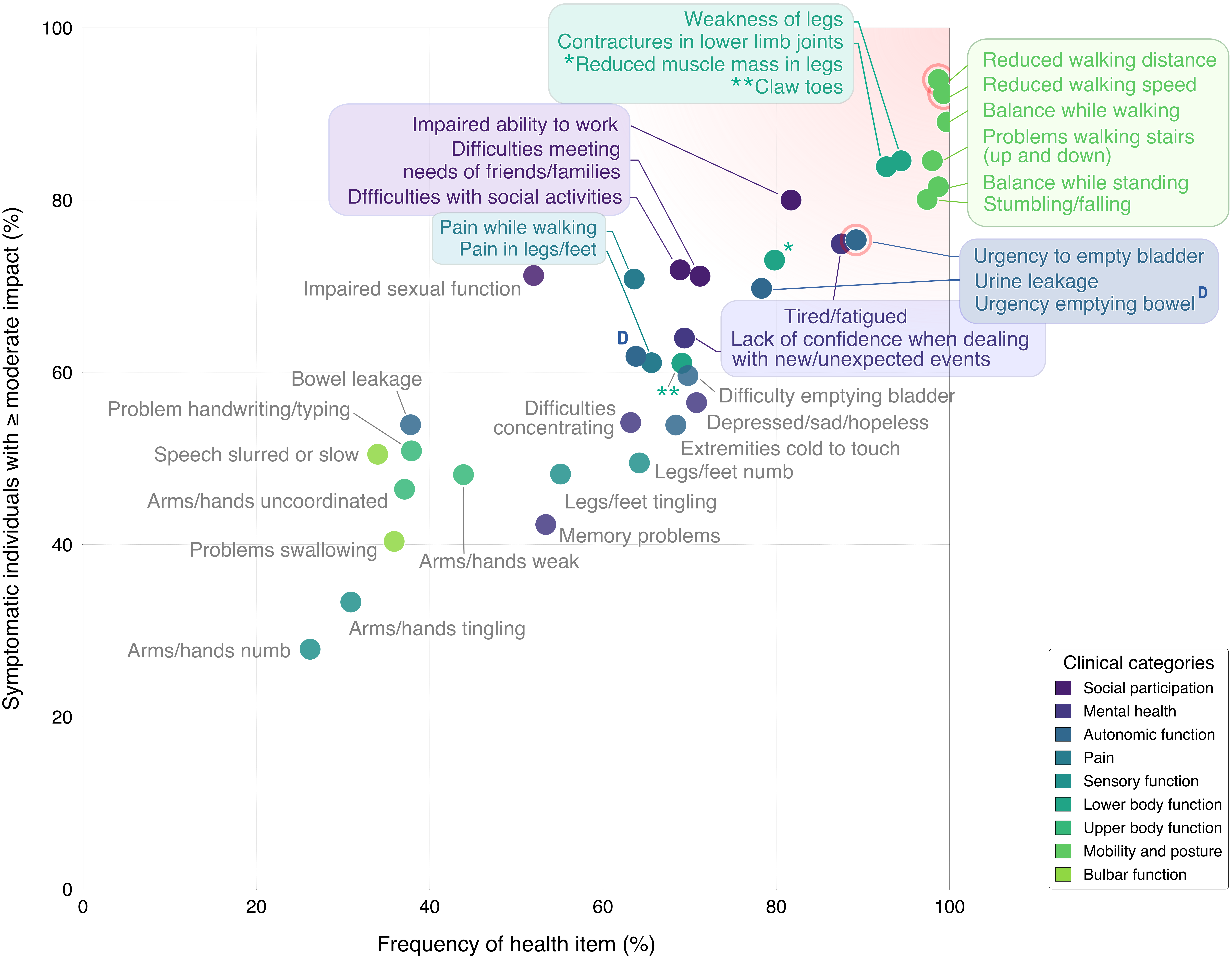


Supplementary Figure 8

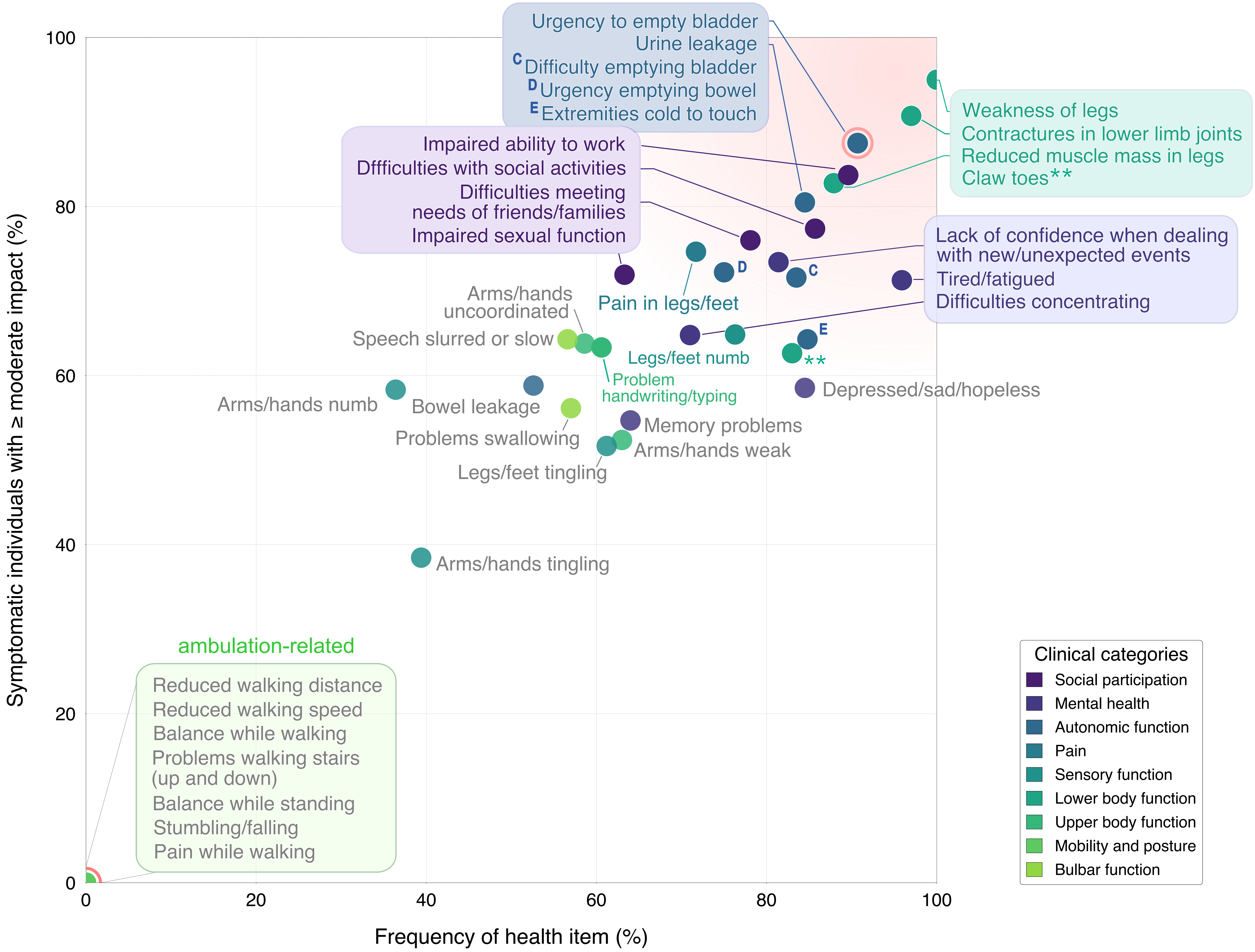


Supplementary Figure 9

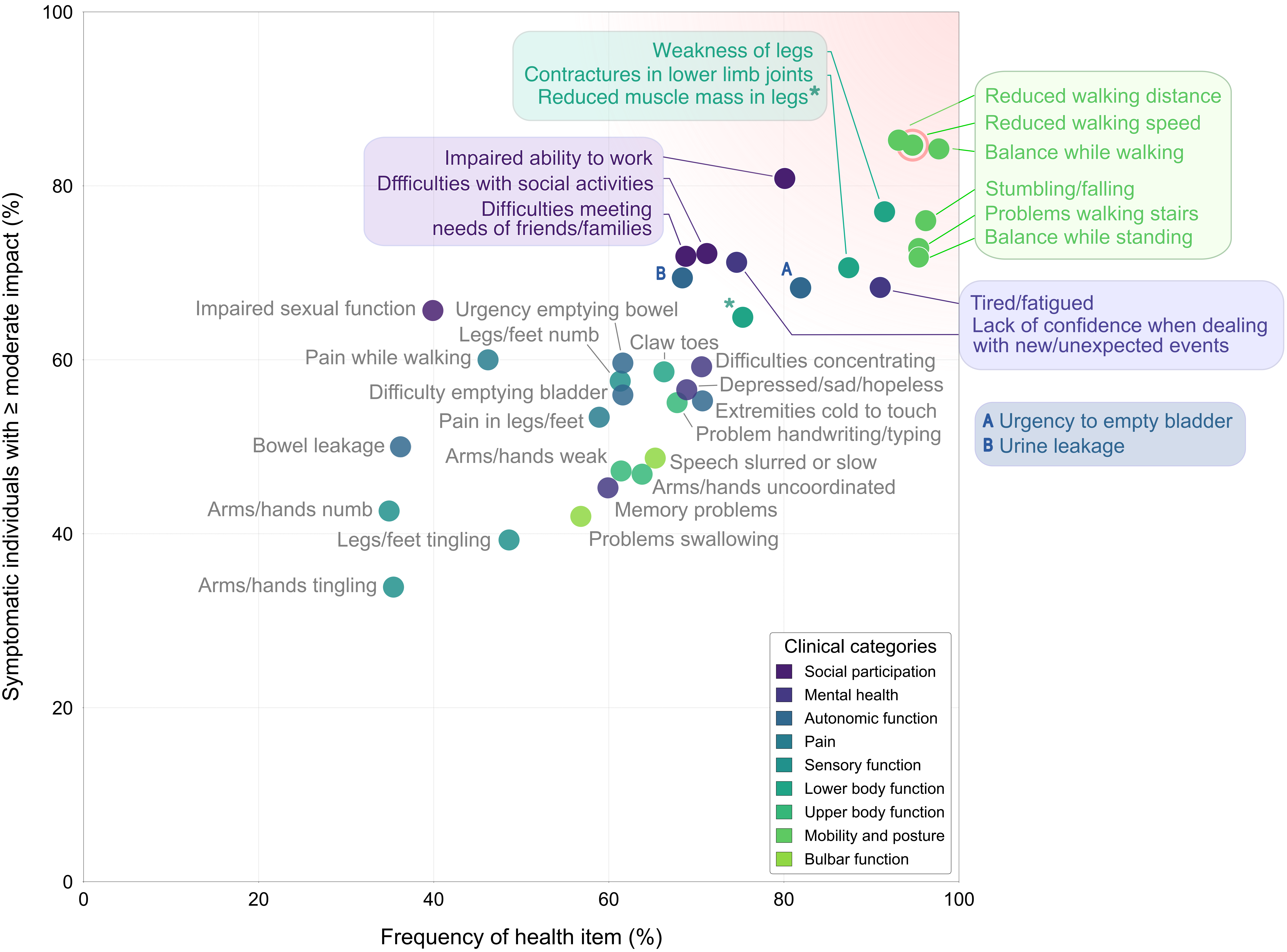


### Supplementary Figure 10
