## Supplementary Tables for "Identifying trial-relevant concepts of interest in HSP: insights from an international patient-voice study in over 600 individuals"

**Supplementary Table S1.** Genotypes of survey respondents in Survey 1 and 2.

| Genotype entry | Survey 1 (n, %) | Survey 2 (n, %) |
|---|---|---|
| HSP – predominant | | |
| GCH1 | 1 (0.2%) | 1 (0.2%) |
| NEFL | 1 (0.2%) | 3 (0.6%) |
| OPTN | 0 (0%) | 1 (0.2%) |
| SPG3 | 9 (1.5%) | 8 (1.6%) |
| SPG4 | 185 (30.0%) | 135 (26.8%) |
| SPG5 | 9 (1.5%) | 8 (1.6%) |
| SPG6 | 1 (0.2%) | 1 (0.2%) |
| SPG8 | 2 (0.3%) | 0 (0%) |
| SPG9 | 3 (0.5%) | 1 (0.2%) |
| SPG10 | 3 (0.5%) | 2 (0.4%) |
| SPG26 | 1 (0.2%) | 0 (0%) |
| SPG30 | 4 (0.6%) | 2 (0.4%) |
| SPG31 | 1 (0.2%) | 5 (1.0%) |
| SPG36 | 1 (0.2%) | 0 (0%) |
| SPG43 | 1 (0.2%) | 0 (0%) |
| SPG50 | 1 (0.2%) | 2 (0.4%) |
| SPG52 | 0 (0%) | 1 (0.2%) |
| SPG56 | 2 (0.3%) | 1 (0.2%) |
| SPG72 | 1 (0.2%) | 0 (0%) |
| SPG76 | 3 (0.5%) | 1 (0.2%) |
| SPG80 | 0 (0%) | 2 (0.4%) |
| SPG81 | 0 (0%) | 0 (0%) |
| SPG83 | 1 (0.2%) | 0 (0%) |
| Unknown | 182 (29.5%) | 152 (30.2%) |
| SPAX subgroup | | |
| AOA2 | 2 (0.3%) | 0 (0%) |
| POLR3A | 4 (0.6%) | 4 (0.8%) |
| ARSACS | 69 (11.2%) | 82 (16.3%) |
| SPG7 | 102 (16.6%) | 72 (14.3%) |
| SPG11 | 21 (3.4%) | 13 (2.6%) |
| SPG15 | 1 (0.2%) | 3 (0.6%) |
| SPG35 | 4 (0.6%) | 1 (0.2%) |
| SPG39 | 1 (0.2%) | 0 (0%) |
| SPG46 | 0 (0%) | 1 (0.2%) |
| SPG49 | 0 (0%) | 1 (0.2%) |
| SPG79 | 0 (0%) | 1 (0.2%) |

**Supplementary Table S2.** Disease stage classification in Survey 1 and 2.

| Survey 1 | Survey 2 | Disease stage classification |
|---|---|---|
| **Select that statement that best describes your mobility:** | **FARS Staging (PROM version)[1)]**<br>**Select the statement that best describes your stage of ataxia or hereditary spastic paraplegia (HSP):** | |
| | ○ Symptoms not present. **(0)** | Pre-symptomatic |
| ○ Never use a walking device or wheelchair | ○ My doctor has detected some minimal signs of my disease during examination. However, I can run or jump without loss of balance. I experience no disability. **(1)**<br>○ I experience mild symptoms of ataxia or HSP. I cannot run or jump without losing balance. I am physically able to lead an independent life, but I am somewhat restricted in my daily activities. I experience minimal disability. **(2)** | Mild |
| ○ Use a stick, walker or other walking device at least some of the time, but never use a wheelchair.<br>○ Use a walking device at least some of the time and a wheelchair at least some of the time. | ○ My ataxic or HSP symptoms are clearly noticeable to others. I often hold onto wall/furniture, another person or use a cane for stability and walking. I experience mild disability. **(3)**<br>○ I require a walker, crutches or two canes for walking or use other walking aids such as a walking dog. I am independent in several activities of daily living. I experience moderate disability. **(4)** | Intermediate |
| ○ Use a wheelchair all the time | ○ I am not able to walk, even with support and use a wheelchair to move about. I am able. to navigate my wheelchair by myself. I can perform some activities of daily living that do not require standing or walking. I experience severe disability. **(5)**<br>○ I am confined to a wheelchair or bed. I completely depend on others for all activities of daily living. **(6)** | Advanced |

[1)] FARS Staging 0: presymptomatic; 0.5 – 2.5: mild; 3.0 – 4.0: intermediate; 4.5 – 6.0: advanced

**Supplementary Table S3**. Prevalence of self-reported health impacts in HSP, grouped by category [Survey 1].

| Categories | Included items | Present at any time over the course of the disease | Currently present (by disease stage) | | | | | Present at any time (SPAX vs. HSP) | | |
|---|---|---|---|---|---|---|---|---|---|---|
| | | Total cohort | Total cohort | Early | Intermediate | Advanced | p-value* | SPAX | HSP (-SPAX) | p-value* |
| Vision and hearing | **≥ 1 symptom from category** | **226/542 41.7%** | | | | | | | | |
| | Reduced visual acuity[3)] | 146/567 25.7% | 110/563 19.5% | 18/143 12.6% | 71/356 19.9% | 21/64 32.8% | **0.00604** | 65/197 33.0% | 81/370 21.9% | **0.01351** |
| | Reduced visual field[2)] | 65/564 11.5% | 38/560 6.8% | 8/144 5.6% | 27/353 7.6% | 3/63 4.8% | 0.55187 | 32/192 16.7% | 33/372 8.9% | **0.01776** |
| | Double vision[1)] | 72/561 12.8% | 37/557 6.6% | 9/143 6.3% | 20/351 5.7% | 8/63 12.7% | 0.19229 | 44/193 22.8% | 28/368 7.6% | **5.48e-06** |
| | Oscillopsia[3)] | 131/567 23.1% | 86/563 15.3% | 19/144 13.2% | 55/353 15.6% | 12/66 18.2% | 0.62562 | 69/195 35.4% | 62/372 16.7% | **6.92e-06** |
| | Hearing impairment[1)] | 97/560 17.3% | 77/556 13.8% | 13/140 9.3% | 53/352 15.1% | 11/64 17.2% | 0.19647 | 39/191 20.4% | 58/369 15.7% | 0.26466 |
| Bulbar function | **≥ 1 symptom from category** | **287/548 52.4%** | | | | | | | | |
| | Drooling / excessive saliva[4)] | 124/564 22.0% | 89/560 15.9% | 15/142 10.6% | 52/353 14.7% | 22/65 33.8% | **0.00023** | 65/195 33.3% | 59/369 16.0% | **2.27e-05** |
| | Slowing of speech[4)] | 173/567 30.5% | 145/564 25.7% | 28/145 19.3% | 92/354 26.0% | 25/65 38.5% | **0.02085** | 104/195 53.3% | 69/372 18.5% | **4.38e-16** |
| | Unable to speak loudly enough[2)] | 89/560 15.9% | 76/557 13.6% | 15/143 10.5% | 45/350 12.9% | 16/64 25.0% | **0.02189** | 43/193 22.3% | 46/367 12.5% | **0.00973** |
| | Slurred speech / dysarthria[4)] | 172/564 30.5% | 138/560 24.6% | 31/142 21.8% | 83/354 23.4% | 24/64 37.5% | **0.04704** | 105/194 54.1% | 67/370 18.1% | **7.43e-17** |
| | Problems with swallowing[4)] | 216/570 37.9% | 159/566 28.1% | 24/141 17.0% | 98/356 27.5% | 37/69 53.6% | **1.03e-06** | 94/194 48.5% | 122/376 32.4% | **0.00099** |
| Mobility and posture | **≥ 1 symptom from category** | **504/504 100%** | | | | | | | | |
| | Reduced walking distance | 576/612 94.1% | 476/608 78.3% | 102/151 67.5% | 345/381 90.6% | 29/76 38.2% | **2.24e-24** | 187/202 92.6% | 389/410 94.9% | 0.3464 |
| | Reduced walking speed | 591/610 96.9% | 495/606 81.7% | 116/151 76.8% | 351/380 92.4% | 28/75 37.3% | **4.05e-27** | 193/203 95.1% | 398/407 97.8% | 0.13395 |
| | Increased exertion while walking[4)] | 567/600 94.5% | 476/596 79.9% | 101/146 69.2% | 344/376 91.5% | 31/74 41.9% | **6.81e-23** | 185/199 93.0% | 382/401 95.3% | 0.34035 |

| | | | | | | | | | | |
|---|---|---|---|---|---|---|---|---|---|---|
| | Dizziness while walking[4)] | 156/563<br>27.7% | 92/560<br>16.4% | 16/142<br>11.3% | 69/354<br>19.5% | 7/64<br>10.9% | **0.04704** | 70/191<br>36.6% | 86/372<br>23.1% | **0.0029** |
| | Problems with balance while walking[4)] | 564/600<br>94.0% | 477/596<br>80.0% | 111/146<br>76.0% | 331/378<br>87.6% | 35/72<br>48.6% | **1.24e-12** | 194/201<br>96.5% | 370/399<br>92.7% | 0.13069 |
| | Problems walking uphill[4)] | 412/568<br>72.5% | 330/564<br>58.5% | 57/143<br>39.9% | 250/356<br>70.2% | 23/65<br>35.4% | **1.00e-11** | 142/196<br>72.4% | 270/372<br>72.6% | 0.97334 |
| | Problems walking downhill[4)] | 541/595<br>90.9% | 444/591<br>75.1% | 102/148<br>68.9% | 318/370<br>85.9% | 24/73<br>32.9% | **1.97e-20** | 177/198<br>89.4% | 364/397<br>91.7% | 0.45175 |
| | Prefer/need handrail to walk up or down stairs | 580/606<br>95.7% | 502/602<br>83.4% | 117/149<br>78.5% | 353/381<br>92.7% | 32/72<br>44.4% | **2.72e-22** | 197/202<br>97.5% | 383/404<br>94.8% | 0.20738 |
| | Stumbling (other than occasional)[4)] | 564/601<br>93.8% | 420/597<br>70.4% | 104/149<br>69.8% | 292/372<br>78.5% | 24/76<br>31.6% | **3.86e-14** | 187/202<br>92.6% | 377/399<br>94.5% | 0.45175 |
| | Falling[4)] | 515/594<br>86.7% | 327/590<br>55.4% | 60/146<br>41.1% | 236/370<br>63.8% | 31/74<br>41.9% | **3.39e-06** | 172/197<br>87.3% | 343/397<br>86.4% | 0.78101 |
| | Dizziness while standing[4)] | 156/560<br>27.9% | 94/557<br>16.9% | 15/144<br>10.4% | 66/349<br>18.9% | 13/64<br>20.3% | 0.06509 | 64/192<br>33.3% | 92/368<br>25.0% | 0.0834 |
| | Problems with balance while standing[4)] | 483/590<br>81.9% | 412/586<br>70.3% | 82/146<br>56.2% | 287/368<br>78.0% | 43/72<br>59.7% | **3.32e-06** | 176/200<br>88.0% | 307/390<br>78.7% | **0.01734** |
| | Scoliosis (abnormal spine curvature)[2)] | 155/561<br>27.6% | 121/557<br>21.7% | 19/142<br>13.4% | 83/348<br>23.9% | 19/67<br>28.4% | **0.02189** | 43/191<br>22.5% | 112/370<br>30.3% | 0.1095 |
| | **≥ 1 symptom from category** | **243/534<br>45.5%** | | | | | | | | |
| | Clumsiness of arms and hands | 216/565<br>38.2% | 179/562<br>31.9% | 40/146<br>27.4% | 108/353<br>30.6% | 31/63<br>49.2% | **0.00995** | 111/193<br>57.5% | 105/372<br>28.2% | **2.48e-10** |
| **Upper body function** | Weakness of arms | 158/568<br>27.8% | 120/564<br>21.3% | 21/146<br>14.4% | 78/351<br>22.2% | 21/67<br>31.3% | **0.02189** | 73/194<br>37.6% | 85/374<br>22.7% | **0.00097** |
| | Muscle wasting upper limbs[1)] | 99/554<br>17.9% | 74/551<br>13.4% | 15/144<br>10.4% | 46/345<br>13.3% | 13/62<br>21.0% | 0.14418 | 34/186<br>18.3% | 65/368<br>17.7% | 0.87084 |
| | Contractures in upper limb joints[1)] | 91/560<br>16.2% | 72/556<br>12.9% | 10/143<br>7.0% | 44/347<br>12.7% | 18/66<br>27.3% | **0.00062** | 24/190<br>12.6% | 67/370<br>18.1% | 0.1769 |
| | **≥ 1 symptom from category** | **506/510<br>99.2%** | | | | | | | | |
| | Feeling of "heavy legs" when walking[4)] | 485/592<br>81.9% | 396/588<br>67.3% | 81/147<br>55.1% | 285/371<br>76.8% | 30/70<br>42.9% | **1.86e-09** | 145/199<br>72.9% | 340/393<br>86.5% | **0.00034** |
| **Lower body function** | Weakness of legs | 528/593<br>89.0% | 451/589<br>76.6% | 106/147<br>72.1% | 304/372<br>81.7% | 41/70<br>58.6% | **0.00016** | 168/197<br>85.3% | 360/396<br>90.9% | 0.08497 |
| | Muscle wasting lower limbs | 373/579<br>64.4% | 321/575<br>55.8% | 54/146<br>37.0% | 218/361<br>60.4% | 49/68<br>72.1% | **8.93e-07** | 112/194<br>57.7% | 261/385<br>67.8% | 0.0424 |
| | Contractures in lower limb joints[4)] | 372/574<br>64.8% | 337/570<br>59.1% | 65/141<br>46.1% | 223/362<br>61.6% | 49/67<br>73.1% | **0.00069** | 102/196<br>52.0% | 270/378<br>71.4% | **3.39e-05** |

| | | | | | | | | | | |
|---|---|---|---|---|---|---|---|---|---|---|
| | Stiffness of muscles[4)] | 491/576 85.2% | 441/572 77.1% | 93/139 66.9% | 290/361 80.3% | 58/72 80.6% | **0.00829** | 161/192 83.9% | 330/384 85.9% | 0.58357 |
| | Muscle cramps[3)] | 301/568 53.0% | 219/565 38.8% | 41/143 28.7% | 138/354 39.0% | 40/68 58.8% | **0.00041** | 104/192 54.2% | 197/376 52.4% | 0.72058 |
| | Claw toes | 277/567 48.9% | 237/563 42.1% | 38/142 26.8% | 170/355 47.9% | 29/66 43.9% | **0.00026** | 89/196 45.4% | 188/371 50.7% | 0.33694 |
| **Sensory function** | **≥ 1 symptom from category** | **400/549 72.9%** | | | | | | | | |
| | Numbness in arms or hands | 157/567 27.7% | 110/563 19.5% | 17/146 11.6% | 71/351 20.2% | 22/66 33.3% | **0.00212** | 70/193 36.3% | 87/374 23.3% | **0.00392** |
| | Tingling in arms or hands | 177/568 31.2% | 117/565 20.7% | 17/143 11.9% | 79/358 22.1% | 21/64 32.8% | **0.00328** | 70/193 36.3% | 107/375 28.5% | 0.12234 |
| | Numbness in legs, feet or toes | 327/576 56.8% | 249/572 43.5% | 48/147 32.7% | 168/361 46.5% | 33/64 51.6% | **0.01072** | 103/193 53.4% | 224/383 58.5% | 0.34035 |
| | Tingling in legs, feet or toes | 350/581 60.2% | 269/577 46.6% | 47/147 32.0% | 184/361 51.0% | 38/69 55.1% | **0.00045** | 97/193 50.3% | 253/388 65.2% | **0.00238** |
| **Pain** | **≥ 1 symptom from category** | **444/527 84.3%** | | | | | | | | |
| | Frequent headache[3)] | 168/564 29.8% | 88/560 15.7% | 13/142 9.2% | 58/353 16.4% | 17/65 26.2% | **0.01072** | 66/194 34.0% | 102/370 27.6% | 0.19938 |
| | Pain in the arms or hands | 120/562 21.4% | 76/559 13.6% | 11/143 7.7% | 51/353 14.4% | 14/63 22.2% | **0.02189** | 44/190 23.2% | 76/372 20.4% | 0.53388 |
| | Pain in the hip or groin area[3)] | 269/574 46.9% | 189/571 33.1% | 33/141 23.4% | 132/363 36.4% | 24/67 35.8% | **0.02653** | 70/195 35.9% | 199/379 52.5% | **0.00097** |
| | Pain in the legs or feet | 344/574 59.9% | 265/571 46.4% | 59/145 40.7% | 179/358 50.0% | 27/68 39.7% | 0.09833 | 106/193 54.9% | 238/381 62.5% | 0.15375 |
| | Regular or persistent neck or shoulder pain[3)] | 254/569 44.6% | 173/565 30.6% | 34/141 24.1% | 109/358 30.4% | 30/66 45.5% | **0.01298** | 81/194 41.8% | 173/375 46.1% | 0.4172 |
| | Regular or persistent lower back pain[3)] | 360/575 62.6% | 268/571 46.9% | 55/147 37.4% | 180/359 50.1% | 33/65 50.8% | **0.0362** | 108/194 55.7% | 252/381 66.1% | **0.03697** |
| | Pain associated with walking[4)] | 342/580 59.0% | 254/576 44.1% | 54/146 37.0% | 177/358 49.4% | 23/72 31.9% | **0.00632** | 88/194 45.4% | 254/386 65.8% | **2.27e-05** |
| | Pain associated with standing[4)] | 253/568 44.5% | 198/564 35.1% | 24/143 16.8% | 153/355 43.1% | 21/66 31.8% | **8.93e-07** | 64/192 33.3% | 189/376 50.3% | **0.00083** |
| **Autonomic function** | **≥ 1 symptom from category** | **379/436 86.9%** | | | | | | | | |
| | Bladder incontinence | 386/592 65.2% | 307/588 52.2% | 51/146 34.9% | 213/369 57.7% | 43/73 58.9% | **3.33e-05** | 127/200 63.5% | 259/392 66.1% | 0.60569 |
| | Difficulty/delay in emptying the bladder | 305/575 53.0% | 245/571 42.9% | 35/143 24.5% | 170/358 47.5% | 40/70 57.1% | **2.87e-06** | 82/192 42.7% | 223/383 58.2% | **0.00213** |

| | | | | | | | | | | |
|---|---|---|---|---|---|---|---|---|---|---|
| | Bowel incontinence | 191/569<br>33.6% | 121/565<br>21.4% | 20/142<br>14.1% | 83/354<br>23.4% | 18/69<br>26.1% | 0.05306 | 62/193<br>32.1% | 129/376<br>34.3% | 0.64917 |
| | Severe constipation[2)] | 179/561<br>31.9% | 105/557<br>18.9% | 16/145<br>11.0% | 67/345<br>19.4% | 22/67<br>32.8% | **0.00167** | 55/187<br>29.4% | 124/374<br>33.2% | 0.45732 |
| | Cold extremities | 360/580<br>62.1% | 291/576<br>50.5% | 53/146<br>36.3% | 188/358<br>52.5% | 50/72<br>69.4% | **4.23e-05** | 119/197<br>60.4% | 241/383<br>62.9% | 0.6117 |
| | Erectile dysfunction (men only) [4)] | 112/275<br>40.7% | 91/272<br>70.2% | 11/72<br>15.3% | 67/170<br>39.4% | 13/30<br>43.3% | **0.00064** | 27/98<br>27.6% | 85/177<br>48.0% | **0.00094** |
| **Metabolism and sleep** | **≥ 1 symptom from category** | **468/519<br>90.2%** | | | | | | | | |
| | Trouble falling asleep[3)] | 256/571<br>44.8% | 179/567<br>31.6% | 33/144<br>22.9% | 124/355<br>34.9% | 22/68<br>32.4% | **0.04224** | 73/197<br>37.1% | 183/374<br>48.9% | **0.01893** |
| | Trouble staying asleep[3)] | 270/567<br>47.6% | 212/563<br>37.7% | 39/142<br>27.5% | 140/354<br>39.5% | 33/67<br>49.3% | **0.00866** | 85/194<br>43.8% | 185/373<br>49.6% | 0.29912 |
| | Being overweight[1)] | 207/558<br>37.1% | 168/554<br>30.3% | 33/143<br>23.1% | 104/343<br>30.3% | 31/68<br>45.6% | **0.00752** | 65/187<br>34.8% | 142/371<br>38.3% | 0.50646 |
| | Being underweight[1)] | 85/548<br>15.5% | 45/545<br>8.3% | 12/140<br>8.6% | 31/343<br>9.0% | 2/62<br>3.2% | 0.33064 | 43/190<br>22.6% | 42/358<br>11.7% | **0.00318** |
| | Tiredness/fatigue/lack of stamina | 479/592<br>80.9% | 416/588<br>70.7% | 87/147<br>59.2% | 287/370<br>77.6% | 42/71<br>59.2% | **4.59e-05** | 170/198<br>85.9% | 309/394<br>78.4% | 0.07019 |
| | Epileptic seizures[1)] | 28/558<br>5.0% | 8/555<br>1.4% | 1/141<br>0.7% | 5/349<br>1.4% | 2/65<br>3.1% | 0.46783 | 13/191<br>6.8% | 15/367<br>4.1% | 0.26466 |
| **Mental health** | **≥ 1 symptom from category** | **437/533<br>82.0%** | | | | | | | | |
| | Feeling depressed/sad/hopeless | 351/574<br>61.1% | 221/570<br>38.6% | 45/143<br>31.5% | 146/360<br>40.2% | 30/67<br>44.8% | 0.11109 | 116/195<br>59.5% | 235/379<br>62.0% | 0.6117 |
| | Regular or persistent feelings of anxiety[3)] | 263/571<br>46.1% | 180/567<br>31.7% | 38/143<br>26.6% | 119/357<br>33.3% | 23/67<br>34.3% | 0.33064 | 85/190<br>44.7% | 178/381<br>46.7% | 0.6952 |
| | Regular or persistent feelings of anger[1)] | 204/561<br>36.4% | 124/557<br>22.3% | 26/142<br>18.3% | 82/354<br>23.2% | 16/61<br>26.2% | 0.39032 | 75/190<br>39.5% | 129/371<br>34.8% | 0.36419 |
| | Lack of attention/concentration | 278/574<br>48.4% | 222/570<br>38.9% | 44/147<br>29.9% | 146/355<br>41.1% | 32/68<br>47.1% | **0.03019** | 109/196<br>55.6% | 169/378<br>44.7% | **0.03587** |
| | Lack of confidence, e.g. when dealing with new or unexpected events | 295/567<br>52.0% | 230/563<br>40.9% | 46/141<br>32.6% | 148/353<br>41.9% | 36/69<br>52.2% | **0.02838** | 106/190<br>55.8% | 189/377<br>50.1% | 0.30696 |
| | Memory problems | 248/574<br>43.2% | 188/570<br>33.0% | 29/143<br>20.3% | 127/357<br>35.6% | 32/70<br>45.7% | **0.00062** | 92/196<br>46.9% | 156/378<br>41.3% | 0.29912 |
| **Social participation** | **≥ 1 symptom from category** | **456/532<br>85.7%** | | | | | | | | |
| | Difficulty with social activities | 313/579<br>54.1% | 255/575<br>44.3% | 57/144<br>39.6% | 167/365<br>45.8% | 31/66<br>47.0% | 0.42523 | 120/197<br>60.9% | 193/382<br>50.5% | **0.0424** |

| | | | | | | | | | | |
|---|---|---|---|---|---|---|---|---|---|---|
| | Reduced ability to work | 381/574 66.4% | 326/570 57.2% | 51/144 35.4% | 230/360 63.9% | 45/66 68.2% | **4.34e-08** | 137/195 70.3% | 244/379 64.4% | 0.26466 |
| | Difficulty meeting needs of friends[4)] | 273/565 48.3% | 221/561 39.4% | 38/143 26.6% | 156/355 43.9% | 27/63 42.9% | **0.00283** | 99/191 51.8% | 174/374 46.5% | 0.33694 |
| | Difficulty meeting needs of family[4)] | 274/565 48.5% | 233/561 41.5% | 33/141 23.4% | 167/351 47.6% | 33/69 47.8% | **1.17e-05** | 96/189 50.8% | 178/376 47.3% | 0.52299 |
| | Reduced interest in sexual activity[4)] | 283/574 49.3% | 232/570 40.7% | 38/143 26.6% | 166/358 46.4% | 28/69 40.6% | **0.00062** | 78/191 40.8% | 205/383 53.5% | **0.01351** |

1) Symptom had high rate of missing data (≥ 8% missing data) and was therefore omitted from Survey 2.
2) Symptom was infrequent (< 30% of the cohort) and was therefore omitted from Survey 2.
3) Symptom considered to be unspecific or unlikely to progress systematically over the course of the disease.
4) Symptoms were partly combined for Survey 2; for details see Material & Methods.

Numbers are N or percentages (%) where indicated. Frequencies are based on valid responses in corresponding sub-item; thus, denominators vary slightly across domains due to missing data.
*P*-values are from Pearson's Chi-square tests followed by Benjamini-Hochberg FDR correction. FDR-adjusted $P < 0.05$ were considered statistically significant and are highlighted in bold.

**Supplementary Table S4.** Prevalence of self-reported health impacts in HSP, grouped by gender [Survey 1].

| Categories | Included symptoms | Present at any time over the course of the disease (male vs. female) | | | | | | | | |
|---|---|---|---|---|---|---|---|---|---|---|
| | | Total cohort | | | HSP (- SPAX) | | | SPAX | | |
| | | Male | Female | p-value* | Male | Female | p-value* | Male | Female | p-value* |
| **Vision and hearing** | | | | | | | | | | |
| | Reduced visual acuity | 58/273 21.2% | 87/288 30.2% | **0.04447** | 27 / 170 (15.9) | 53 / 195 (27.2) | **0.035** | 31 / 103 (30.1) | 34 / 93 (36.6) | 0.615 |
| | Reduced visual field | 27/271 10.0% | 38/287 13.2% | 0.34167 | 11 / 171 (6.4) | 22 / 196 (11.2) | 0.2157 | 16 / 100 (16.0) | 16 / 91 (17.6) | 0.8569 |
| | Double vision | 34/271 12.5% | 38/284 13.4% | 0.84148 | 12 / 170 (7.1) | 16 / 193 (8.3) | 0.7354 | 22 / 101 (21.8) | 22 / 91 (24.2) | 0.8396 |
| | Oscillopsia | 52/273 19.0% | 77/288 26.7% | 0.06797 | 22 / 171 (12.9) | 39 / 196 (19.9) | 0.169 | 30 / 102 (29.4) | 38 / 92 (41.3) | 0.2964 |
| | Hearing impairment | 52/270 19.3% | 45/284 15.8% | 0.41767 | 28 / 169 (16.6) | 30 / 195 (15.4) | 0.8105 | 24 / 101 (23.8) | 15 / 89 (16.9) | 0.5162 |
| **Bulbar function** | | | | | | | | | | |
| | Drooling / excessive saliva | 68/271 25.1% | 55/287 19.2% | 0.16648 | 28 / 169 (16.6) | 31 / 195 (15.9) | 0.8883 | 40 / 102 (39.2) | 24 / 92 (26.1) | 0.2398 |
| | Slowing of speech | 100/272 36.8% | 71/289 24.6% | **0.00904** | 37 / 170 (21.8) | 31 / 197 (15.7) | 0.2281 | 63 / 102 (61.8) | 40 / 92 (43.5) | 0.1399 |
| | Unable to speak loudly enough | 49/269 18.2% | 39/285 13.7% | 0.24185 | 21 / 168 (12.5) | 25 / 194 (12.9) | 0.9257 | 28 / 101 (27.7) | 14 / 91 (15.4) | 0.2066 |
| | Slurred speech / dysarthria | 100/270 37.0% | 71/288 24.7% | **0.00873** | 36 / 168 (21.4) | 31 / 197 (15.7) | 0.2591 | 64 / 102 (62.7) | 40 / 91 (44.0) | 0.1399 |
| | Problems with swallowing | 102/274 37.2% | 111/290 38.3% | 0.84626 | 51 / 173 (29.5) | 69 / 198 (34.8) | 0.3804 | 51 / 101 (50.5) | 42 / 92 (45.7) | 0.7318 |
| **Mobility and posture** | | | | | | | | | | |
| | Reduced walking distance | 275/291 94.5% | 295/314 93.9% | 0.84148 | 179 / 186 (96.2) | 205 / 218 (94.0) | 0.4273 | 96 / 105 (91.4) | 90 / 96 (93.8) | 0.7318 |
| | Reduced walking speed | 283/289 97.9% | 301/314 95.9% | 0.24185 | 181 / 184 (98.4) | 211 / 217 (97.2) | 0.5479 | 102 / 105 (97.1) | 90 / 97 (92.8) | 0.3785 |
| | Increased exertion while walking | 273/287 95.1% | 287/306 93.8% | 0.61308 | 174 / 183 (95.1) | 202 / 212 (95.3) | 0.9258 | 99 / 104 (95.2) | 85 / 94 (90.4) | 0.4259 |
| | Dizziness while walking | 56/266 21.1% | 99/291 34.0% | **0.00446** | 27 / 168 (16.1) | 58 / 199 (29.1) | **0.0233** | 29 / 98 (29.6) | 41 / 92 (44.6) | 0.1868 |
| | Problems with balance while walking | 262/282 92.9% | 296/312 94.9% | 0.44562 | 162 / 179 (90.5) | 203 / 215 (94.4) | 0.2281 | 100 / 103 (97.1) | 93 / 97 (95.9) | 0.8195 |

| | | | | | | | | | | |
|---|---|---|---|---|---|---|---|---|---|---|
| | Problems walking uphill | 184/269<br>68.4% | 224/292<br>76.7% | 0.06481 | 111 / 168<br>(66.1) | 155 / 198<br>(78.3) | **0.035** | 73 / 101<br>(72.3) | 69 / 94<br>(73.4) | 0.8945 |
| | Problems walking downhill | 261/284<br>91.9% | 274/304<br>90.1% | 0.59098 | 167 / 181<br>(92.3) | 192 / 210<br>(91.4) | 0.8105 | 94 / 103<br>(91.3) | 82 / 94<br>(87.2) | 0.6213 |
| | Prefer/need handrail to walk up or down stairs | 274/288<br>95.1% | 299/311<br>96.1% | 0.66967 | 171 / 184<br>(92.9) | 206 / 214<br>(96.3) | 0.2281 | 103 / 104<br>(99.0) | 93 / 97<br>(95.9) | 0.3785 |
| | Stumbling (other than occasional) | 267/286<br>93.4% | 290/308<br>94.2% | 0.77729 | 170 / 182<br>(93.4) | 201 / 211<br>(95.3) | 0.5434 | 97 / 104<br>(93.3) | 89 / 97<br>(91.8) | 0.8396 |
| | Falling | 235/279<br>84.2% | 274/308<br>89.0% | 0.16648 | 147 / 179<br>(82.1) | 191 / 212<br>(90.1) | 0.0683 | 88 / 100<br>(88.0) | 83 / 96<br>(86.5) | 0.8569 |
| | Dizziness while standing | 54/267<br>20.2% | 101/287<br>35.2% | **0.00079** | 29 / 168<br>(17.3) | 62 / 195<br>(31.8) | **0.0166** | 25 / 99<br>(25.3) | 39 / 92<br>(42.4) | 0.1399 |
| | Problems with balance while standing | 219/279<br>78.5% | 259/305<br>84.9% | 0.09542 | 126 / 175<br>(72.0) | 177 / 210<br>(84.3) | **0.0233** | 93 / 104<br>(89.4) | 82 / 95<br>(86.3) | 0.7318 |
| | Scoliosis (abnormal spine curvature) | 62/271<br>22.9% | 91/284<br>32.0% | **0.04447** | 45 / 171<br>(26.3) | 65 / 194<br>(33.5) | 0.2281 | 17 / 100<br>(17.0) | 26 / 90<br>(28.9) | 0.2398 |
| **Upper body function** | | | | | | | | | | |
| | Clumsiness of arms and hands | 101/270<br>37.4% | 113/289<br>39.1% | 0.77729 | 40 / 169<br>(23.7) | 64 / 198<br>(32.3) | 0.169 | 61 / 101<br>(60.4) | 49 / 91<br>(53.8) | 0.6213 |
| | Weakness of arms | 60/270<br>22.2% | 96/292<br>32.9% | **0.01778** | 30 / 170<br>(17.6) | 54 / 199<br>(27.1) | 0.087 | 30 / 100<br>(30.0) | 42 / 93<br>(45.2) | 0.1868 |
| | Muscle wasting upper limbs | 47/267<br>17.6% | 50/280<br>17.9% | 0.9380 | 27 / 170<br>(15.9) | 36 / 192<br>(18.8) | 0.5721 | 20 / 97<br>(20.6) | 14 / 88<br>(15.9) | 0.656 |
| | Contractures in upper limb joints | 41/269<br>15.2% | 48/284<br>16.9% | 0.70052 | 27 / 169<br>(16.0) | 38 / 195<br>(19.5) | 0.4987 | 14 / 100<br>(14.0) | 10 / 89<br>(11.2) | 0.7458 |
| **Lower body function** | | | | | | | | | | |
| | Feeling of "heavy legs" when walking | 221/283<br>78.1% | 259/302<br>85.8% | **0.04447** | 150 / 179<br>(83.8) | 186 / 208<br>(89.4) | 0.2089 | 71 / 104<br>(68.3) | 73 / 94<br>(77.7) | 0.3785 |
| | Weakness of legs | 243/279<br>87.1% | 278/307<br>90.6% | 0.28733 | 155 / 177<br>(87.6) | 199 / 213<br>(93.4) | 0.1238 | 88 / 102<br>(86.3) | 79 / 94<br>(84.0) | 0.8282 |
| | Muscle wasting lower limbs | 170/279<br>60.9% | 197/293<br>67.2% | 0.20022 | 112 / 177<br>(63.3) | 144 / 202<br>(71.3) | 0.2019 | 58 / 102<br>(56.9) | 53 / 91<br>(58.2) | 0.8945 |
| | Contractures in lower limb joints | 171/271<br>63.1% | 197/296<br>66.6% | 0.5267 | 116 / 169<br>(68.6) | 151 / 203<br>(74.4) | 0.3234 | 55 / 102<br>(53.9) | 46 / 93<br>(49.5) | 0.7318 |
| | Stiffness of muscles | 228/275<br>82.9% | 258/296<br>87.2% | 0.24662 | 145 / 174<br>(83.3) | 181 / 206<br>(87.9) | 0.3183 | 83 / 101<br>(82.2) | 77 / 90<br>(85.6) | 0.7318 |
| | Muscle cramps | 139/271<br>51.3% | 156/290<br>53.8% | 0.66967 | 85 / 169<br>(50.3) | 107 / 201<br>(53.2) | 0.6595 | 54 / 102<br>(52.9) | 49 / 89<br>(55.1) | 0.8569 |
| | Claw toes | 117/269<br>43.5% | 156/291<br>53.6% | **0.04447** | 72 / 167<br>(43.1) | 113 / 198<br>(57.1) | **0.035** | 45 / 102<br>(44.1) | 43 / 93<br>(46.2) | 0.8569 |
| **Sensory function** | | | | | | | | | | |

| | | | | | | | | | | |
|---|---|---|---|---|---|---|---|---|---|---|
| | Numbness in arms or hands | 74/272<br>27.2% | 81/289<br>28.0% | 0.86541 | 38 / 170<br>(22.4) | 48 / 199<br>(24.1) | 0.7545 | 36 / 102<br>(35.3) | 33 / 90<br>(36.7) | 0.8945 |
| | Tingling in arms or hands | 75/274<br>27.4% | 98/287<br>34.1% | 0.15803 | 44 / 173<br>(25.4) | 60 / 196<br>(30.6) | 0.3804 | 31 / 101<br>(30.7) | 38 / 91<br>(41.8) | 0.3301 |
| | Numbness in legs, feet or toes | 139/275<br>50.5% | 183/294<br>62.2% | **0.01778** | 93 / 174<br>(53.4) | 126 / 203<br>(62.1) | 0.1958 | 46 / 101<br>(45.5) | 57 / 91<br>(62.6) | 0.1528 |
| | Tingling in legs, feet or toes | 144/278<br>51.8% | 202/297<br>68.0% | **0.00079** | 104 / 178<br>(58.4) | 146 / 205<br>(71.2) | **0.035** | 40 / 100<br>(40.0) | 56 / 92<br>(60.9) | 0.1332 |
| **Pain** | | | | | | | | | | |
| | Frequent headache | 63/271<br>23.2% | 101/287<br>35.2% | **0.00904** | 34 / 169<br>(20.1) | 65 / 196<br>(33.2) | **0.0298** | 29 / 102<br>(28.4) | 36 / 91<br>(39.6) | 0.3301 |
| | Pain in the arms or hands | 43/267<br>16.1% | 73/288<br>25.3% | **0.02451** | 26 / 169<br>(15.4) | 47 / 197<br>(23.9) | 0.119 | 17 / 98<br>(17.3) | 26 / 91<br>(28.6) | 0.2674 |
| | Pain in the hip or groin area | 108/270<br>40.0% | 156/297<br>52.5% | **0.01218** | 76 / 170<br>(44.7) | 119 / 203<br>(58.6) | **0.035** | 32 / 100<br>(32.0) | 37 / 94<br>(39.4) | 0.5699 |
| | Pain in the legs or feet | 150/272<br>55.1% | 189/295<br>64.1% | 0.06797 | 99 / 172<br>(57.6) | 135 / 203<br>(66.5) | 0.172 | 51 / 100<br>(51.0) | 54 / 92<br>(58.7) | 0.5699 |
| | Regular or persistent neck or shoulder pain | 93/271<br>34.3% | 155/291<br>53.3% | **0.00014** | 58 / 171<br>(33.9) | 110 / 198<br>(55.6) | **0.0005** | 35 / 100<br>(35.0) | 45 / 93<br>(48.4) | 0.2555 |
| | Regular or persistent lower back pain | 148/270<br>54.8% | 206/298<br>69.1% | **0.00337** | 98 / 170<br>(57.6) | 149 / 205<br>(72.7) | **0.0193** | 50 / 100<br>(50.0) | 57 / 93<br>(61.3) | 0.3301 |
| | Pain associated with walking | 146/273<br>53.5% | 190/300<br>63.3% | **0.04447** | 107 / 173<br>(61.8) | 142 / 207<br>(68.6) | 0.2635 | 39 / 100<br>(39.0) | 48 / 93<br>(51.6) | 0.2964 |
| | Pain associated with standing | 103/271<br>38.0% | 145/290<br>50.0% | **0.0173** | 72 / 171<br>(42.1) | 113 / 199<br>(56.8) | **0.0298** | 31 / 100<br>(31.0) | 32 / 91<br>(35.2) | 0.7318 |
| **Autonomic function** | | | | | | | | | | |
| | Bladder incontinence | 176/281<br>62.6% | 206/304<br>67.8% | 0.2957 | 111 / 178<br>(62.4) | 145 / 208<br>(69.7) | 0.2281 | 65 / 103<br>(63.1) | 61 / 96<br>(63.5) | 0.9608 |
| | Difficulty/delay in emptying the bladder | 159/279<br>57.0% | 144/290<br>49.7% | 0.15699 | 116 / 178<br>(65.2) | 106 / 200<br>(53.0) | 0.0541 | 43 / 101<br>(42.6) | 38 / 90<br>(42.2) | 0.9608 |
| | Bowel incontinence | 91/274<br>33.2% | 97/288<br>33.7% | 0.91959 | 56 / 172<br>(32.6) | 71 / 198<br>(35.9) | 0.6006 | 35 / 102<br>(34.3) | 26 / 90<br>(28.9) | 0.6593 |
| | Severe constipation | 76/271<br>28.0% | 101/285<br>35.4% | 0.12831 | 54 / 171<br>(31.6) | 68 / 198<br>(34.3) | 0.6595 | 22 / 100<br>(22.0) | 33 / 87<br>(37.9) | 0.1528 |
| | Erectile dysfunction (men only) | 112/275<br>40.7% | NA | NA | 85 / 177<br>(48.0) | 0 / 121<br>(0.0) | NA | 27 / 98<br>(27.6) | 0 / 61<br>(0.0) | NA |
| | Cold extremities | 133/273<br>48.7% | 222/300<br>74.0% | **0.00000** | 81 / 172<br>(47.1) | 156 / 205<br>(76.1) | **0.0000** | 52 / 101<br>(51.5) | 66 / 95<br>(69.5) | 0.1399 |
| **Metabolism and sleep** | | | | | | | | | | |
| | Trouble falling asleep | 98/277<br>35.4% | 154/287<br>53.7% | **0.00022** | 65 / 174<br>(37.4) | 115 / 194<br>(59.3) | **0.0005** | 33 / 103<br>(32.0) | 39 / 93<br>(41.9) | 0.3785 |

| | | | | | | | | | | |
|---|---|---|---|---|---|---|---|---|---|---|
| | Trouble staying asleep | 111/276 40.2% | 154/284 54.2% | **0.00566** | 73 / 173 (42.2) | 107 / 194 (55.2) | **0.0455** | 38 / 103 (36.9) | 47 / 90 (52.2) | 0.1868 |
| | Being overweight | 88/273 32.2% | 116/279 41.6% | 0.05785 | 58 / 173 (33.5) | 82 / 193 (42.5) | 0.1741 | 30 / 100 (30.0) | 34 / 86 (39.5) | 0.41 |
| | Being underweight | 39/265 14.7% | 46/277 16.6% | 0.66967 | 16 / 166 (9.6) | 26 / 187 (13.9) | 0.3234 | 23 / 99 (23.2) | 20 / 90 (22.2) | 0.8945 |
| | Tiredness/fatigue/lack of stamina | 215/282 76.2% | 258/303 85.1% | **0.02145** | 129 / 178 (72.5) | 175 / 210 (83.3) | **0.035** | 86 / 104 (82.7) | 83 / 93 (89.2) | 0.4259 |
| | Epileptic seizures | 15/269 5.6% | 13/283 4.6% | 0.70052 | 10 / 169 (5.9) | 5 / 193 (2.6) | 0.2168 | 5 / 100 (5.0) | 8 / 90 (8.9) | 0.5699 |
| **Mental health** | | | | | | | | | | |
| | Feeling depressed/sad/hopeless | 144/274 52.6% | 203/294 69.0% | **0.00077** | 90 / 173 (52.0) | 142 / 201 (70.6) | **0.003** | 54 / 101 (53.5) | 61 / 93 (65.6) | 0.2964 |
| | Regular or persistent feelings of anxiety | 106/273 38.8% | 151/291 51.9% | **0.00904** | 67 / 173 (38.7) | 106 / 202 (52.5) | **0.035** | 39 / 100 (39.0) | 45 / 89 (50.6) | 0.3301 |
| | Regular or persistent feelings of anger | 93/270 34.4% | 108/284 38.0% | 0.52515 | 55 / 170 (32.4) | 72 / 195 (36.9) | 0.4784 | 38 / 100 (38.0) | 36 / 89 (40.4) | 0.8569 |
| | Lack of attention/concentration | 131/275 47.6% | 143/293 48.8% | 0.84148 | 72 / 172 (41.9) | 94 / 201 (46.8) | 0.4626 | 59 / 103 (57.3) | 49 / 92 (53.3) | 0.7458 |
| | Lack of confidence, e.g. when dealing with new or unexpected events | 129/268 48.1% | 163/293 55.6% | 0.15388 | 78 / 170 (45.9) | 109 / 202 (54.0) | 0.2249 | 51 / 98 (52.0) | 54 / 91 (59.3) | 0.5998 |
| | Memory problems | 118/275 42.9% | 128/293 43.7% | 0.87729 | 69 / 172 (40.1) | 86 / 201 (42.8) | 0.6809 | 49 / 103 (47.6) | 42 / 92 (45.7) | 0.8635 |
| **Social participation** | | | | | | | | | | |
| | Difficulty with social activities | 138/275 50.2% | 170/298 57.0% | 0.17626 | 77 / 171 (45.0) | 112 / 206 (54.4) | 0.169 | 61 / 104 (58.7) | 58 / 92 (63.0) | 0.7318 |
| | Reduced ability to work | 178/276 64.5% | 201/292 68.8% | 0.39971 | 109 / 174 (62.6) | 133 / 200 (66.5) | 0.5474 | 69 / 102 (67.6) | 68 / 92 (73.9) | 0.615 |
| | Difficulty meeting needs of friends | 136/272 50.0% | 134/287 46.7% | 0.57556 | 81 / 172 (47.1) | 90 / 197 (45.7) | 0.8225 | 55 / 100 (55.0) | 44 / 90 (48.9) | 0.656 |
| | Difficulty meeting needs of family | 118/271 43.5% | 153/288 53.1% | 0.05785 | 71 / 173 (41.0) | 104 / 198 (52.5) | 0.0812 | 47 / 98 (48.0) | 49 / 90 (54.4) | 0.6298 |
| | Reduced interest in sexual activity | 114/276 41.3% | 168/291 57.7% | **0.00079** | 81 / 177 (45.8) | 123 / 200 (61.5) | **0.0193** | 33 / 99 (33.3) | 45 / 91 (49.5) | 0.1845 |

**Supplementary Table S5:** Severity and relevance of health-related impacts in daily life.

| Included items | Severity | | | | | | | Relevance[1] | | | | | |
|---|---|---|---|---|---|---|---|---|---|---|---|---|---|
| | Median/ IQR | Not present | Mild | Moderate | Severe | Very severe | Missing responses | Median/ IQR | No effect | Small effect | Moderate effect | Severe effect | Very severe effect |
| **Bulbar function** | | | | | | | | | | | | | |
| Speech slurred or slow | 0.0 (0.0 - 1.0) | 316 / 501 (63.1%) | 97 / 501 (19.4%) | 52 / 501 (10.4%) | 28 / 501 (5.6%) | 8 / 501 (1.6%) | 3 / 504 (0.6%) | 2.0 (1.0 - 2.0) | 12 / 185 (6.5%) | 74 / 185 (40.0%) | 53 / 185 (28.6%) | 35 / 185 (18.9%) | 11 / 185 (5.9%) |
| Problem swallowing | 0.0 (0.0 - 1.0) | 315 / 503 (62.6%) | 105 / 503 (20.9%) | 55 / 503 (10.9%) | 23 / 503 (4.6%) | 5 / 503 (1.0%) | 1 / 504 (0.2%) | 1.0 (1.0-2.0) | 16 / 188 (8.5%) | 90 / 188 (47.9%) | 54 / 188 (28.7%) | 21 / 188 (11.2%) | 7 / 188 (3.7%) |
| **Mobility and posture** | | | | | | | | | | | | | |
| Reduced walking distance | 3.0 (2.0 - 3.0) | 14 / 391 (3.6%) | 34 / 391 (8.7%) | 94 / 391 (24.0%) | 178 / 391 (45.5%) | 71 / 391 (18.2%) | 113 / 504 (22.4%) | 3.0 (2.0 - 3.0) | 9 / 377 (2.4%) | 39 / 377 (10.3%) | 126 / 377 (33.4%) | 158 / 377 (41.9%) | 45 / 377 (11.9%) |
| Reduced walking speed | 3.0 (2.0 - 3.0) | 11 / 391 (2.8%) | 25 / 391 (6.4%) | 110 / 391 (28.1%) | 175 / 391 (44.8%) | 70 / 391 (17.9%) | 113 / 504 (22.4%) | 3.0 (2.0 - 3.0) | 11 / 380 (2.9%) | 37 / 380 (9.7%) | 133 / 380 (35.0%) | 150 / 380 (39.5%) | 49 / 380 (12.9%) |
| Balance while walking | 2.0 (2.0 - 3.0) | 13 / 391 (3.3%) | 69 / 391 (17.6%) | 121 / 391 (30.9%) | 142 / 391 (36.3%) | 46 / 391 (11.8%) | 113 / 504 (22.4%) | 2.0 (2.0 - 3.0) | 13 / 378 (3.4%) | 58 / 378 (15.3%) | 136 / 378 (36.0%) | 132 / 378 (34.9%) | 39 / 378 (10.3%) |
| Balance while standing | 2.0 (1.0 - 3.0) | 29 / 391 (7.4%) | 102 / 391 (26.1%) | 137 / 391 (35.0%) | 100 / 391 (25.6%) | 23 / 391 (5.9%) | 113 / 504 (22.4%) | 2.0 (1.0 - 3.0) | 11 / 361 (3.0%) | 92 / 361 (25.5%) | 141 / 361 (39.1%) | 96 / 361 (26.6%) | 21 / 361 (5.8%) |
| Stumbling/falling | 2.0 (1.0 - 2.0) | 14 / 389 (3.6%) | 105 / 389 (27.0%) | 173 / 389 (44.5%) | 81 / 389 (20.8%) | 16 / 389 (4.1%) | 115 / 504 (22.8%) | 2.0 (2.0 - 3.0) | 4 / 374 (1.1%) | 87 / 374 (23.3%) | 157 / 374 (42.0%) | 110 / 374 (29.4%) | 16 / 374 (4.3%) |
| Problems walking stairs | 2.0 (2.0 - 3.0) | 14 / 392 (3.6%) | 63 / 392 (16.1%) | 132 / 392 (33.7%) | 133 / 392 (33.9%) | 50 / 392 (12.8%) | 112 / 504 (22.2%) | 2.0 (2.0 - 3.0) | 8 / 377 (2.1%) | 79 / 377 (21.0%) | 130 / 377 (34.5%) | 117 / 377 (31.0%) | 43 / 377 (11.4%) |
| **Upper body function** | | | | | | | | | | | | | |
| Arms/hands weak | 0.0 (0.0 - 1.0) | 270 / 502 (53.8%) | 136 / 502 (27.1%) | 78 / 502 (15.5%) | 14 / 502 (2.8%) | 4 / 502 (0.8%) | 2 / 504 (0.4%) | 1.0 (1.0 - 2.0) | 18 / 232 (7.8%) | 106 / 232 (45.7%) | 77 / 232 (33.2%) | 25 / 232 (10.8%) | 6 / 232 (2.6%) |
| Arms/hands uncoordinated | 0.0 (0.0 - 1.0) | 303 / 500 (60.6%) | 120 / 500 (24.0%) | 58 / 500 (11.6%) | 17 / 500 (3.4%) | 2 / 500 (0.4%) | 4 / 504 (0.8%) | 1.0 (1.0 - 2.0) | 12 / 195 (6.2%) | 87 / 195 (44.6%) | 67 / 195 (34.4%) | 25 / 195 (12.8%) | 4 / 195 (2.1%) |
| Problem handwriting/typing | 0.0 (0.0 - 1.0) | 293 / 499 (58.7%) | 106 / 499 (21.2%) | 64 / 499 (12.8%) | 30 / 499 (6.0%) | 6 / 499 (1.2%) | 5 / 504 (1.0%) | 2.0 (1.0 - 2.0) | 15 / 205 (7.3%) | 84 / 205 (41.0%) | 72 / 205 (35.1%) | 27 / 205 (13.2%) | 7 / 205 (3.4%) |
| **Lower body function** | | | | | | | | | | | | | |
| Leg weakness | 2.0 (2.0 - 3.0) | 30 / 500 (6.0%) | 81 / 500 (16.2%) | 179 / 500 (35.8%) | 147 / 500 (29.4%) | 63 / 500 (12.6%) | 4 / 504 (0.8%) | 2.0 (2.0 - 3.0) | 11 / 469 (2.3%) | 70 / 469 (14.9%) | 169 / 469 (36.0%) | 153 / 469 (32.6%) | 66 / 469 (14.1%) |
| Lower limb contractures | 2.0 (1.0 - 3.0) | 42 / 500 (8.4%) | 93 / 500 (18.6%) | 176 / 500 (35.2%) | 142 / 500 (28.4%) | 47 / 500 (9.4%) | 4 / 504 (0.8%) | 2.0 (2.0 - 3.0) | 12 / 458 (2.6%) | 79 / 458 (17.2%) | 176 / 458 (38.4%) | 134 / 458 (29.3%) | 57 / 458 (12.4%) |
| Muscle wasting in legs | 2.0 (1.0 - 2.0) | 108 / 500 (21.6%) | 133 / 500 (26.6%) | 147 / 500 (29.4%) | 83 / 500 (16.6%) | 29 / 500 (5.8%) | 4 / 504 (0.8%) | 2.0 (1.0 - 3.0) | 19 / 391 (4.9%) | 94 / 391 (24.0%) | 157 / 391 (40.2%) | 89 / 391 (22.8%) | 32 / 391 (8.2%) |
| Claw toes | 1.0 (0.0 - 2.0) | 167 / 500 (33.4%) | 98 / 500 (19.6%) | 119 / 500 (23.8%) | 78 / 500 (15.6%) | 38 / 500 (7.6%) | 4 / 504 (0.8%) | 2.0 (1.0 - 2.0) | 40 / 331 (12.1%) | 93 / 331 (28.1%) | 116 / 331 (35.0%) | 58 / 331 (17.5%) | 24 / 331 (7.3%) |
| **Sensory function** | | | | | | | | | | | | | |
| Legs/feet numb | 1.0 (0.0 - 2.0) | 185 / 498 (37.1%) | 150 / 498 (30.1%) | 106 / 498 (21.3%) | 45 / 498 (9.0%) | 12 / 498 (2.4%) | 6 / 504 (1.2%) | 2.0 (1.0 - 2.0) | 25 / 312 (8.0%) | 128 / 312 (41.0%) | 92 / 312 (29.5%) | 52 / 312 (16.7%) | 15 / 312 (4.8%) |
| Legs/feet tingling | 1.0 (0.0 - 2.0) | 223 / 498 (44.8%) | 132 / 498 (26.5%) | 93 / 498 (18.7%) | 44 / 498 (8.8%) | 6 / 498 (1.2%) | 6 / 504 (1.2%) | 1.0 (1.0 - 2.0) | 33 / 274 (12.0%) | 109 / 274 (39.8%) | 81 / 274 (29.6%) | 42 / 274 (15.3%) | 9 / 274 (3.3%) |
| Arms/hands numb | 0.0 (0.0 - 1.0) | 365 / 499 (73.1%) | 90 / 499 (18.0%) | 38 / 499 (7.6%) | 3 / 499 (0.6%) | 3 / 499 (0.6%) | 5 / 504 (1.0%) | 1.0 (1.0 - 2.0) | 10 / 134 (7.5%) | 76 / 134 (56.7%) | 40 / 134 (29.9%) | 5 / 134 (3.7%) | 3 / 134 (2.2%) |
| Arms/hands tingling | 0.0 (0.0 - 1.0) | 346 / 498 (69.5%) | 98 / 498 (19.7%) | 45 / 498 (9.0%) | 6 / 498 (1.2%) | 3 / 498 (0.6%) | 6 / 504 (1.2%) | 1.0 (1.0 - 2.0) | 17 / 152 (11.2%) | 85 / 152 (55.9%) | 40 / 152 (26.3%) | 7 / 152 (4.6%) | 3 / 152 (2.0%) |
| **Pain** | | | | | | | | | | | | | |
| Pain in legs/feet | 1.0 (0.0 - 2.0) | 168 / 500 (33.6%) | 126 / 500 (25.2%) | 116 / 500 (23.2%) | 69 / 500 (13.8%) | 21 / 500 (4.2%) | 4 / 504 (0.8%) | 2.0 (1.0 - 3.0) | 14 / 331 (4.2%) | 111 / 331 (33.5%) | 102 / 331 (30.8%) | 70 / 331 (21.1%) | 34 / 331 (10.3%) |
| Pain while walking | 1.0 (0.0 - 2.0) | 149 / 390 (38.2%) | 112 / 390 (28.7%) | 78 / 390 (20.0%) | 38 / 390 (9.7%) | 13 / 390 (3.3%) | 114 / 504 (22.6%) | 2.0 (1.0 - 3.0) | 5 / 241 (2.1%) | 79 / 241 (32.8%) | 83 / 241 (34.4%) | 58 / 241 (24.1%) | 16 / 241 (6.6%) |
| **Autonomic function** | | | | | | | | | | | | | |

| | | | | | | | | | | | | | |
|---|---|---|---|---|---|---|---|---|---|---|---|---|---|
| Urgency to empty bladder | 3.0 (1.0 - 3.0) | 71 / 500 (14.2%) | 55 / 500 (11.0%) | 110 / 500 (22.0%) | 204 / 500 (40.8%) | 60 / 500 (12.0%) | 4 / 504 (0.8%) | 2.0 (2.0 - 3.0) | 28 / 428 (6.5%) | 73 / 428 (17.1%) | 133 / 428 (31.1%) | 127 / 428 (29.7%) | 67 / 428 (15.7%) |
| Urine leakage | 2.0 (0.0 - 3.0) | 125 / 498 (25.1%) | 92 / 498 (18.5%) | 132 / 498 (26.5%) | 100 / 498 (20.1%) | 49 / 498 (9.8%) | 6 / 504 (1.2%) | 2.0 (1.0 - 3.0) | 23 / 373 (6.2%) | 83 / 373 (22.3%) | 117 / 373 (31.4%) | 99 / 373 (26.5%) | 51 / 373 (13.7%) |
| Difficulty emptying bladder | 2.0 (0.0 - 3.0) | 154 / 499 (30.9%) | 94 / 499 (18.8%) | 111 / 499 (22.2%) | 95 / 499 (19.0%) | 45 / 499 (9.0%) | 5 / 504 (1.0%) | 2.0 (1.0 - 3.0) | 38 / 341 (11.1%) | 93 / 341 (27.3%) | 113 / 341 (33.1%) | 56 / 341 (16.4%) | 41 / 341 (12.0%) |
| Extremities cold to touch | 1.0 (0.0 - 2.0) | 157 / 498 (31.5%) | 110 / 498 (22.1%) | 123 / 498 (24.7%) | 83 / 498 (16.7%) | 25 / 498 (5.0%) | 6 / 504 (1.2%) | 2.0 (1.0 - 2.0) | 59 / 340 (17.4%) | 93 / 340 (27.4%) | 126 / 340 (37.1%) | 45 / 340 (13.2%) | 17 / 340 (5.0%) |
| Urgency emptying bowel | 1.0 (0.0 - 2.0) | 188 / 498 (37.8%) | 93 / 498 (18.7%) | 140 / 498 (28.1%) | 66 / 498 (13.3%) | 11 / 498 (2.2%) | 6 / 504 (1.2%) | 2.0 (1.0 - 2.0) | 30 / 310 (9.7%) | 94 / 310 (30.3%) | 111 / 310 (35.8%) | 47 / 310 (15.2%) | 28 / 310 (9.0%) |
| Bowel leakage | 0.0 (0.0 - 1.0) | 311 / 499 (62.3%) | 92 / 499 (18.4%) | 70 / 499 (14.0%) | 20 / 499 (4.0%) | 6 / 499 (1.2%) | 5 / 504 (1.0%) | 2.0 (1.0 - 2.0) | 11 / 187 (5.9%) | 71 / 187 (38.0%) | 52 / 187 (27.8%) | 32 / 187 (17.1%) | 21 / 187 (11.2%) |
| **Mental health** | | | | | | | | | | | | | |
| Fatigue | 2.0 (1.0 - 3.0) | 59 / 502 (11.8%) | 116 / 502 (23.1%) | 177 / 502 (35.3%) | 110 / 502 (21.9%) | 40 / 502 (8.0%) | 2 / 504 (0.4%) | 2.0 (1.0 - 3.0) | 13 / 440 (3.0%) | 113 / 440 (25.7%) | 164 / 440 (37.3%) | 103 / 440 (23.4%) | 47 / 440 (10.7%) |
| Poor concentration | 1.0 (0.0 - 2.0) | 181 / 503 (36.0%) | 147 / 503 (29.2%) | 127 / 503 (25.2%) | 41 / 503 (8.2%) | 7 / 503 (1.4%) | 1 / 504 (0.2%) | 2.0 (1.0 - 2.0) | 13 / 320 (4.1%) | 124 / 320 (38.8%) | 124 / 320 (38.8%) | 44 / 320 (13.8%) | 15 / 320 (4.7%) |
| Memory problems | 1.0 (0.0 - 1.0) | 227 / 504 (45.0%) | 167 / 504 (33.1%) | 71 / 504 (14.1%) | 33 / 504 (6.5%) | 6 / 504 (1.2%) | 0 / 504 (0.0%) | 1.0 (1.0 - 2.0) | 22 / 277 (7.9%) | 135 / 277 (48.7%) | 71 / 277 (25.6%) | 35 / 277 (12.6%) | 14 / 277 (5.1%) |
| Depressed/sad/hopeless | 1.0 (0.0 - 2.0) | 147 / 500 (29.4%) | 187 / 500 (37.4%) | 99 / 500 (19.8%) | 44 / 500 (8.8%) | 23 / 500 (4.6%) | 4 / 504 (0.8%) | 2.0 (1.0 - 2.0) | 15 / 352 (4.3%) | 138 / 352 (39.2%) | 122 / 352 (34.7%) | 49 / 352 (13.9%) | 28 / 352 (8.0%) |
| Lack of confidence | 1.0 (0.0 - 2.0) | 153 / 498 (30.7%) | 123 / 498 (24.7%) | 129 / 498 (25.9%) | 60 / 498 (12.0%) | 33 / 498 (6.6%) | 6 / 504 (1.2%) | 2.0 (1.0 - 3.0) | 14 / 345 (4.1%) | 105 / 345 (30.4%) | 129 / 345 (37.4%) | 60 / 345 (17.4%) | 37 / 345 (10.7%) |
| **Social participation** | | | | | | | | | | | | | |
| Impaired ability to work | 2.0 (1.0 - 3.0) | 109 / 495 (22.0%) | 83 / 495 (16.8%) | 96 / 495 (19.4%) | 114 / 495 (23.0%) | 93 / 495 (18.8%) | 9 / 504 (1.8%) | 2.0 (2.0 - 3.0) | 15 / 384 (3.9%) | 63 / 384 (16.4%) | 121 / 384 (31.5%) | 94 / 384 (24.5%) | 91 / 384 (23.7%) |
| Impaired sexual function | 1.0 (0.0 - 2.0) | 238 / 480 (49.6%) | 62 / 480 (12.9%) | 70 / 480 (14.6%) | 71 / 480 (14.8%) | 39 / 480 (8.1%) | 24 / 504 (4.8%) | 2.0 (1.0 - 3.0) | 13 / 241 (5.4%) | 56 / 241 (23.2%) | 65 / 241 (27.0%) | 69 / 241 (28.6%) | 38 / 241 (15.8%) |
| Dffficulties with social activities | 1.0 (0.0 - 2.0) | 153 / 500 (30.6%) | 123 / 500 (24.6%) | 116 / 500 (23.2%) | 72 / 500 (14.4%) | 36 / 500 (7.2%) | 4 / 504 (0.8%) | 2.0 (1.0 - 3.0) | 7 / 347 (2.0%) | 92 / 347 (26.5%) | 135 / 347 (38.9%) | 71 / 347 (20.5%) | 42 / 347 (12.1%) |
| Difficulties meeting needs of friends/families | 1.0 (0.0 - 2.0) | 158 / 495 (31.9%) | 106 / 495 (21.4%) | 122 / 495 (24.6%) | 69 / 495 (13.9%) | 40 / 495 (8.1%) | 9 / 504 (1.8%) | 2.0 (1.0 - 3.0) | 8 / 336 (2.4%) | 93 / 336 (27.7%) | 120 / 336 (35.7%) | 72 / 336 (21.4%) | 43 / 336 (12.8%) |

1) Relevance among patients currently experiencing this symptom.

Number and percent of total are given (N [%])

**Supplementary Table S6.** Polychoric correlation between disease severity and relevance. (limitation: Q1-6 severe stage NaNs)

| | Correlation coefficient |
|---|---|
| **Bulbar function** | |
| Speech slurred or slow | 0.92896 |
| Problem swallowing | 0.91649 |
| **Mobility and Posture** | |
| Reduced walking distance | 0.75120 |
| Reduced walking speed | 0.77066 |
| Balance while walking | 0.86632 |
| Balance while standing | 0.90434 |
| Stumbling/falling | 0.79433 |
| Problems walking stairs | 0.81318 |
| **Upper body function** | |
| Arms/hands weak | 0.90655 |
| Arms/hands uncoordinated | 0.83877 |
| Problem handwriting/typing | 0.94605 |
| **Lower body function** | |
| Leg weakness | 0.84945 |
| Lower limb contractures | 0.86968 |
| Muscle wasting in legs | 0.79322 |
| Claw toes | 0.76498 |
| **Sensory function** | |
| Arms/hands numb | 0.85984 |
| Legs/feet numb | 0.84045 |
| Arms/hands tingling | 0.75033 |
| Legs/feet tingling | 0.82320 |
| **Pain** | |
| Pain in legs/feet | 0.92104 |
| Pain while walking | 0.94014 |
| **Autonomic function** | |
| Urgency to empty bladder | 0.82960 |
| Urine leakage | 0.83264 |
| Difficulty emptying bladder | 0.85426 |
| Urgency emptying bowel | 0.80318 |
| Extremities cold to touch | 0.82788 |
| Bowel leakage | 0.71964 |
| **Mental health** | |
| Fatigue | 0.92873 |
| Poor concentration | 0.87246 |
| Memory problems | 0.91769 |
| Depressed/sad/hopeless | 0.90543 |
| Lack of confidence | 0.93694 |
| **Social participation** | |
| Impaired ability to work | 0.95075 |
| Impaired sexual function | 0.63882 |
| Difficulties with social activities | 0.92852 |
| Difficulties meeting needs of friends/families | 0.87682 |
